# Subpopulation-specific Machine Learning Prognosis for Underrepresented Patients with Double Prioritized Bias Correction

**DOI:** 10.1101/2021.03.26.21254401

**Authors:** Sharmin Afrose, Wenjia Song, Charles B. Nemeroff, Chang Lu, Danfeng (Daphne) Yao

## Abstract

**Background:** Many clinical datasets are intrinsically imbalanced, dominated by overwhelming majority groups. Off-the-shelf machine learning models that optimize the prognosis of majority patient types (e.g., healthy class) may cause substantial errors on the minority prediction class (e.g., disease class) and demographic subgroups (e.g., Black or young patients). In the typical one-machine-learning-model-fits-all paradigm, racial and age disparities are likely to exist, but unreported. In addition, some widely used whole-population metrics give misleading results.

**Methods:** We design a double prioritized (DP) bias correction technique to mitigate representational biases in machine learning-based prognosis. Our method trains customized machine learning models for specific ethnicity or age groups, a substantial departure from the one-model-predicts-all convention. We compare with other sampling and reweighting techniques in mortality and cancer survivability prediction tasks.

**Results:** We first provide empirical evidence showing various prediction deficiencies in a typical machine learning setting without bias correction. For example, missed death cases are 3.14 times higher than missed survival cases for mortality prediction. Then, we show DP consistently boosts the minority class recall for underrepresented groups, by up to 38.0%. DP also reduces relative disparities across race and age groups, e.g., up to 88.0% better than the 8 existing sampling solutions in terms of the relative disparity of minority class recall. Cross-race and cross-age-group evaluation also suggests the need for subpopulation-specific machine learning models.

**Conclusions:** Biases exist in the widely accepted one-machine-learning-model-fits-all-population approach. We invent a bias correction method that produces specialized machine learning prognostication models for underrepresented racial and age groups. This technique may reduce life-threatening prediction mistakes for minority populations.

**Plain Language Summary:** This work aims to improve the prediction accuracy of machine learning models in medical applications, e.g., estimating the likelihood of a patient dying in an emergency room visit or surviving cancer. Inaccurate prediction may produce life-threatening consequences. We first examine how biases in training data impact prediction outcomes, in particular how underrepresented patients (e.g., young patients or patients of color) are impacted. Then, we design a double prioritized (DP) bias correction technique. It allows one to train machine learning models for specific demographic groups, e.g., one machine learning model for Black patients and another model for Asian patients. Our results confirm the need for training subpopulation-specific machine learning models. Our work helps improve the medical care of minority patients in the age of digital health.

## Introduction

Researchers have trained machine learning models to predict many diseases and conditions, including Alzheimer’s disease^1^, heart disease^2^, risk of developing diabetic retinopathy^3^, cancer risk^4^ and survivability^5^, genetic testing for diseases^6^, hypertrophic cardiomyopathy diagnosis^7^, psychosis^8^, posttraumatic stress disorder (PTSD)^9^, and COVID–19^10^. Neural network-powered automatic image analysis has also been shown useful for fast disease detection, e.g., breast cancer^11^ and lung cancer^12^. A study showed that deep learning algorithms diagnose breast cancer more accurately (AUC=0.994) than 11 pathologists^11^. Hospitals (e.g., Cleveland Clinic partnering with Microsoft^13^, Johns Hopkins Hospital partnering with GE Healthcare)^14^ are reported to use predictive analytics for monitoring patients’ health status and preventing emergencies^15–18^.

However, clinical datasets are intrinsically imbalanced due to the naturally occurring frequencies of data^19^. The data is not evenly distributed across prediction classes (e.g., disease class vs. healthy class), race, age, or other subgroups. Data imbalance is a major cause of biased prediction results^19^. Biased prediction results may have serious consequences for some patients. For example, a recent study showed that automatic enrollment of high–risk patients into the health program favors white patients, although Black patients had 26.3% more chronic health conditions than equally ranked white patients^20^. Similarly, algorithmic osteoarthritis pain prediction shows 43% racial disparities^21^. The design of widely used case-control studies is shown to have a temporal bias that reduces predictive accuracy^22^. For non–medical applications, researchers also identified serious biases in high–profile machine learning applications, e.g., a widely deployed recidivism prediction tool^23–25^, online advertisement system^26^, Amazon’s recruiting engine^27^, and face recognition system^28^. The lack of external validation and overclaiming causal effect in machine learning also raise concerns^29^.

A widely used bias-correction approach to the data imbalance problem is sampling. Oversampling, e.g., replicated oversampling (ROS), is to balance a dataset by adding samples of the minority class; undersampling, e.g., random undersampling (RUS), is to balance a dataset by removing samples of the majority class^30^. An improvement is the K–nearest neighbor (K–NN) classifier–based undersampling technique^31^ (e.g., NearMiss1, NearMiss2, NearMiss3, Distant) that selects samples from the majority class based on distance from minority class samples. State-of-the-art solutions are all oversampling methods, including Synthetic Minority Over-sampling Technique (SMOTE)^32^, Adaptive Synthetic Sampling (ADASYN)^33^, and Gamma^34^. All three methods generate new minority points based on existing minority samples, namely using linear interpolation^32^, gamma distribution^34^, or at the class border^33^. However, existing sampling techniques are not designed to address subgroup biases, as they sample the entire minority class. These methods do not differentiate demographic subgroups (e.g., Black patients or young patients under 30). Thus, it is unclear how well existing sampling solutions reduce accuracy disparity.

We present two categories of contributions to machine learning prognosis for underrepresented patients. One contribution is empirical evidence showing severe racial and age prediction disparities and the deceptive nature of some common metrics. Another contribution is on evaluating the bias-correction ability of sampling methods, including a new double prioritized (DP) bias correction technique. In our first contribution, we use two large medical datasets (MIMIC III and SEER) to show multiple types of prediction deficiencies, some due to the choice of metrics. Poor prediction performance in minority samples is not reflected in widely used whole-population metrics. For imbalanced datasets, conventional metrics such as overall accuracy and AUC–ROC are largely influenced by the performance of the majority samples, which machine learning models aim to fit. Unfortunately, this serious deficiency is not well discussed or reported by medical literature. For example, a study showed that 66.7% of the 33 medical-related machine learning papers used AUC–ROC to evaluate models trained on imbalanced datasets^35^. In our second contribution, we present a new technique, double prioritized (DP) bias correction, that aims to improve the prediction accuracy of specific demographic groups through sample enrichment. DP trains customized prediction models for specific subpopulations, a departure from the existing one-model-predicts-all-demographics paradigm. DP prioritizes specific underrepresented groups, as opposed to sampling across the entire patient population.

From our experiments, we report racial, age, and metric disparities in machine learning models trained on clinical prediction benchmark^17^ on MIMIC III and cancer survival prediction^5^ on SEER cancer dataset. Both training datasets are imbalanced in terms of race and age distributions. For example, for the in-hospital mortality (IHM) prediction with MIMIC III, 70.6% of data represents White patients, whereas only 9.6% represents Black patients. MIMIC III and SEER also have data imbalance problems among the two class labels (e.g., death vs. survival).

For the IHM prediction, only 13.5% of data belongs to the patient who died in the hospital. These data imbalances result in serious prediction biases. A typical neural network-based machine learning model^17^ that we tested correctly predicts 87.6% of non-death cases, but only 60.9% of death cases. Meanwhile, overall accuracy (computed over all patients) is relatively high (0.85), and AUC–ROC is 0.86, because of the good performance in the majority class. These high scores are misleading. Our study also reveals that accuracy among age or race subgroups differs. For example, the mortality prediction precision (i.e., the fraction of actual deaths among predicted deaths) of young patients under 30 is 0.09, substantially lower than the whole population (0.40). Recognizing these accuracy challenges will help advance AI-based technologies to better serve underrepresented patients. Our results show that DP is effective in boosting the minority class recall for underrepresented groups, by up to 38.0%. DP also reduces the disparity among age and race groups. For the in-hospital mortality (IHM) and 5-year breast cancer survivability (BCS) predictions, DP shows a 14.8% to 23.9% improvement over the original model and 5.6% to 88.0% improvement over eight existing sampling techniques for the relative disparity of minority class recall. Our cross-race and cross-age-group results also suggest the need for training specialized machine learning models for different demographic subgroups. All sampling techniques (including DP) are not designed to address biases caused by underdiagnosis, measurement, or any other sources of disparity besides data representation. In what follows, DP assumes that the noise is the same across all demographic subgroups and the only source of bias that it aims to correct is representational.

## Methods

### Double prioritized (DP) bias correction method

DP prioritizes a specific demographic subgroup (e.g., Black patients) that suffers from data imbalance by replicating minority prediction class (C1) cases from this group (e.g., Black in-hospital deaths). DP incrementally increases the number of duplicated units and chooses the optimal unit number based on the resulting models’ performance. Figure 1 shows the machine learning workflow with DP bias correction. The main steps are described next.

**Figure 1:**
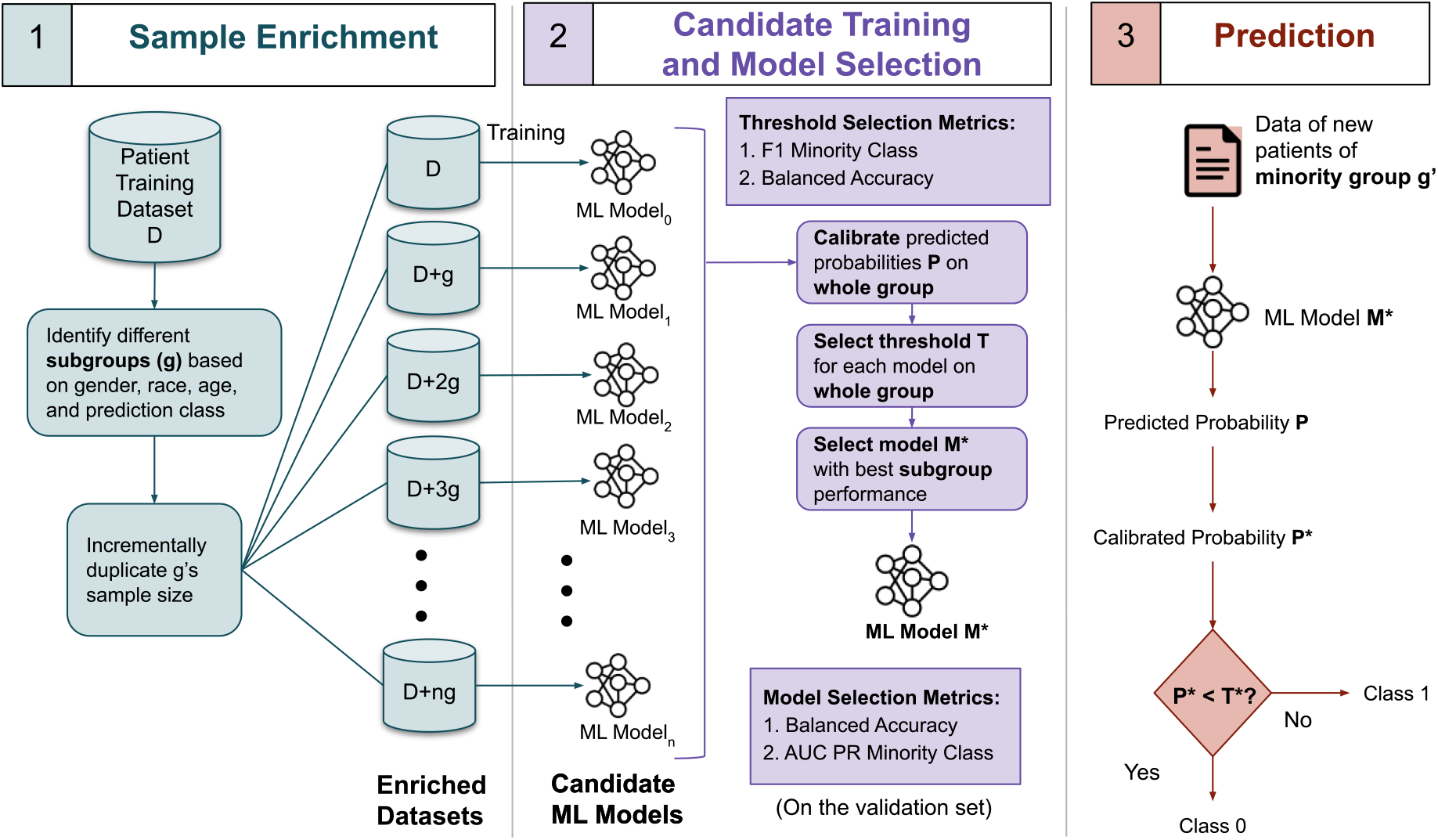
Workflow for improving data balance in machine learning prognosis prediction using double prioritized (DP) bias correction. *Sample enrichment* prepares a number of new training datasets by incrementally enriching a specific demographic subgroup; *candidate training* is where each of the *n*+1 datasets is used for training a candidate machine learning model; *model selection* identifies the optimal model; *prediction* applies the selected model on new patient data. AUC-PR represents the area under the curve of the precision-recall curve.

Sample Enrichment replicates minority class C1 samples in the training dataset for a target demographic group *g* up to *n* times. Each time, duplicated samples are merged with the original training dataset, which forms a new training dataset. Thus, we obtain *n*+1 sets of training datasets, including the original one. Our experiment sets *n* to 19. The value *n* can be empirically determined based on prediction performance.

Candidate Training is to generate a set of candidate machine learning models. Each of the *n*+1 datasets is used to train and generate a candidate machine learning model. Two types of neural networks are used, the long short-term memory (LSTM) model and the multilayer perceptron (MLP) model. Following Harutyunyan *et al*,^17^ for the hospital record prediction tasks, patients’ data is preprocessed into time-series records and fed into an LSTM model. Cancer survivability prediction utilizes an MLP model, following Hegselmann *et al*.^5^ Prediction and data analysis code is in Python programming language. The hospital record prediction tasks were executed on a virtual machine with Ubuntu 18.04 operating system, x86-64 architecture, 8 cores, 40 GB RAM, and 1 GPU. Cancer survivability prediction tasks were performed using an Ubuntu 21.04 operating system, x86-64 architecture, 16 cores, 40 GB RAM, and 1 GPU. Model parameters remain constant in different bias correction techniques (Supplementary Table 1).

Model Selection is to identify the optimal machine learning model among the *n*+1 candidate models. We choose a final machine learning model *M\** after evaluating all candidate models’ performance as follows. For each model, we first calibrate the predicted probabilities on the validation set. Calibration is to adjust the distribution of probabilities before mapping probabilities into labels. We calibrate the output probabilities using the Isotonic Regression technique. We then perform threshold tuning to find the optimal threshold based on balanced accuracy and the F1_C1 score. Specifically, we first identify the top three thresholds that give the highest F1_C1 scores and then further select the optimal threshold that gives the highest balanced accuracy for all samples. For some subgroups, there are only a couple of hundreds of samples in the validation set. Selecting the threshold based on subgroup data may cause overfitting to the validation set. Therefore, we choose thresholds based on the whole group performances. Given a threshold, we then identify the top three machine learning models with the highest balanced accuracy (i.e., average recall of both C0 and C1 classes, Supplementary Equation 6) values and select the model that gives the highest PR_C1 (the area under the curve (AUC) of minority class C1’s precision-recall curve, denoted by AUC-PR_C1 or PR_C1) for demographic group *g*. In this step, no enrichment is applied to the validation dataset. When deciding thresholds, AUC-PR cannot be used, as it is a threshold-free metric. Thus, we use balanced accuracy and F1_C1.

Prediction applies model *M*\* to new patients’ records of minority group *g*’ and obtains a binary class label. At deployment, the demographic group *g* of duplicated samples during Sample enrichment and test group *g*’ should be the same, e.g., the DP model trained with duplicated Black samples is used to predict new Black patients. Evaluation metrics include accuracy, balanced accuracy, Matthews Correlation Coefficient (MCC), AUC–ROC score, precision, recall, AUC–PR, and F1 score of minority and majority prediction classes, the whole population, and various demographic subgroups, including gender (male, female), race (White, Black, Hispanic, Asian), and 8 age groups. Minority class C1 precision calculates the fraction of actual minority C1 class cases among predicted ones. C1 recall calculates the fraction of C1 cases that are successfully predicted by a machine learning model. We use the relative disparity metric to capture the disparity among race groups or age groups. Equation 1 shows the equation for the relative disparity. All other metrics are defined in supplementary equations.

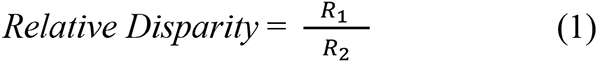

where *R_1_* is the highest and *R_2_* is the lowest evaluation metric value being compared. Similar to other studies^36, 37^, our workflow does not sample the test dataset, because the ground truth (i.e., new patient’s disease or health label) is unknown in the real world. Relative disparity values are greater than or equal to 1. MCC values are in the range of [-1, 1]. The other metric values are in the range of [0, 1]. When comparing datasets that have different percentages of minority class C1 samples, we avoid metrics (e.g., AUC–PR) whose baselines (i.e., the performance of a random classifier) depend on the C1 percentage^35^.

### Other bias correction techniques being compared

The eight existing sampling approaches being compared include four undersampling techniques (namely, random undersampling, NearMiss1, NearMiss3, distant method), and four oversampling techniques (namely, replicated oversampling, SMOTE, ADASYN, Gamma). Undersampling balances the distribution of the two prediction classes by selecting only a subset of the majority class cases. Oversampling balances the dataset by populating the minority class. We also use MLP models with different structures (i.e., different number of layers, different neurons per layer, and different dropout rates).

Reweighting is an alternative bias correction approach to sampling^38, 39^. The reweighting approach assigns different importance to samples in the training data, in order for some minority class samples to impact more on training outcomes. We compare DP with two methods, the standard reweighting method and a new prioritized reweighting method. Standard reweighting aims to make the weights of the two prediction classes balanced. In the standard reweighting approach, new weights are applied to the entire class population as follows. Reweight all samples so that each majority sample weights less than 1 and each minority sample weights more than 1, while satisfying the constraint that the total weight of each prediction class is equal. In our standard reweighting experiment, the minority class has a weight of 3.94 and the majority class has a weight of 0.57 for BCS prediction. The weights are 3.12 and 0.60 for the minority and majority classes, respectively for LCS prediction.

### Prioritized reweighting

Following our DP design, we also invent a new prioritized reweighting approach. Prioritized reweighting selectively reweights specific subgroup minority samples, as opposed to reweighting all minority class C1 samples as in the standard reweighting. In the new prioritized reweighting method, we dynamically reweight minority class samples of selected demographic subgroups and choose the optimal machine learning model using the same metrics and procedure as in DP. Specifically, in each round of prioritized reweighting experiments, we multiply the selected samples’ default weight by a unit number *n*, where *n* ranges from 1 to 20. The weights of samples in other subgroups and majority class samples in the selected subgroup remain the default value, i.e., 1. These weights are used to train a machine learning model. Once the *n* machine learning models are trained, we follow DP’s Model Selection operation for calibration and threshold selection.

### Cross-racial-group and cross-age-group experiments

We also perform a series of cross-group experiments, where enriched samples and test samples are from different demographic groups, i.e., group *g* used for Sample enrichment and test group *g*’ are different. The purpose is to assess the impact of different machine learning models on prediction outcomes.

### Whole-group vs. subgroup-based threshold tuning

When analyzing the performance of the original model without bias correction, we evaluate two different settings. The first setting is to select an optimal threshold based on all samples in the validation set. We refer to the selected threshold as the whole group threshold. The second setting is to select an optimal threshold for each demographic subgroup based on that specific subgroup’s performance in the validation set. We refer to the selected thresholds as the subgroup thresholds. In both settings, we calibrate the prediction on all samples (i.e., whole group) and select the thresholds with the top 3 highest F1 C1 scores and choose the one with the best balanced accuracy.

### SHAP-sum and SHAP-avg feature importance

We calculate the feature importance for all four tasks (i.e., IHM, Decompensation, BCS, and LCS) using the Shapley Additive exPlanations (SHAP). For one-hot encoded categorical variables, each of them is represented by multiple columns in the input data. SHAP is not designed for such one-hot encoded categorical features. The standard SHAP method calculates the importance of each column. Thus, we have to post-process the importance of these features. We implement two approaches, SHAP-avg and SHAP-sum. In the SHAP-avg approach, we compute the average importance of columns representing the same feature, i.e., the importance of columns representing the same variable is averaged. In the SHAP-sum approach, we add up the importance of all columns representing the same feature.

### Clinical datasets

We use MIMIC III^17, 40^ and SEER^41^ cancer datasets, both collected in the US. We test existing machine learning models in a clinical prediction benchmark^17^ for MIMIC III and cancer survival prediction^5^ for SEER. We study a total of four binary classification tasks, in-hospital mortality (IHM) prediction and decompensation prediction from the clinical prediction benchmark,^17^ 5-year breast cancer survivability (BCS) prediction, and 5-year lung cancer survivability (LCS) prediction. In what follows, we denote the minority prediction class as Class 1 (or C1) and the majority class as Class 0 (or C0).

Figure 2a-d shows the composition of IHM training data, which contains 14,681 time-series samples from MIMIC III. The majority of the records (86.5%) belong to Class 0 (i.e., patients who do not die in hospital). The rest (13.5%) belong to Class 1 (i.e., the patients who die in the hospital). The percentage of Class 1 samples within each subgroup slightly varies but is consistently low. 70.6% of the patients are White and 76% belong to the age range [50, 90). 45.1% of the patients are females and 54.9% are males. The training set contains insufficient data for the young adult population. Distributions of the decompensation training dataset (of size 2,377,768) are similar (Supplementary Figure 1a-d). Figure 2e-h shows the percentages of different subgroup sizes for the training dataset used in BCS prediction. The BCS training set contains 199,000 samples, of which 87.3% are in Class 0 (i.e., patients diagnosed with breast cancer and survived for more than 5 years) and 0.6% are males. The percentage of Class 1 samples is low in most groups, with an exception of the age 90+ subgroup, which has a high mortality rate. The majority race group (81%) is White. When categorized by age, 70% of the patients are between 40 and 70. The LCS training dataset (of size 164,443) follows similar imbalanced distributions (Supplementary Figure 1e-h).

**Figure 2:**
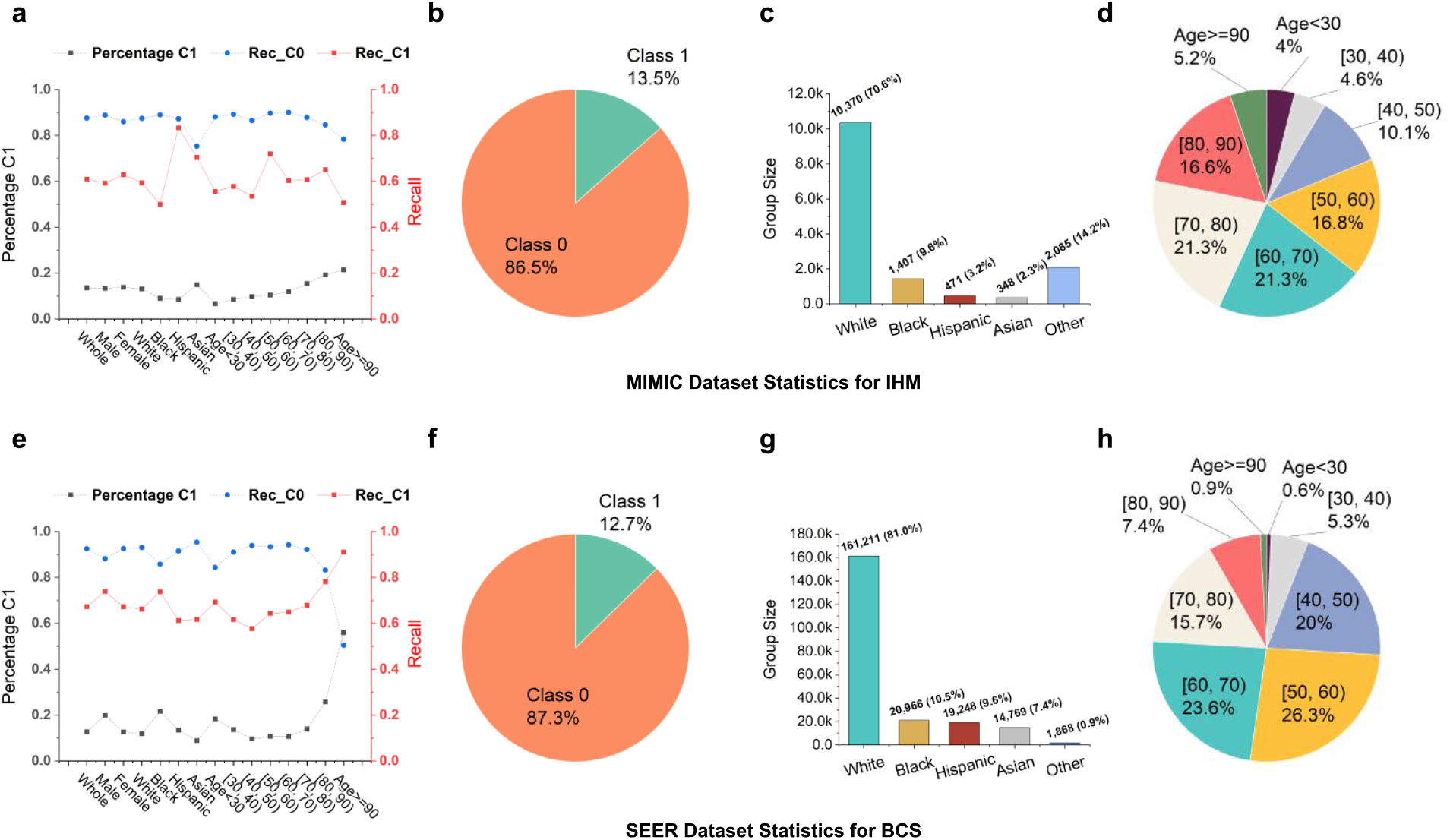
Recall values for both classes C0 and C1 and training data statistics for the in-hospital mortality (IHM) and the 5-year breast cancer survivability (BCS) tasks. **(a)** Percentage of the minority class C1, Recall C0, and Recall C1 of each subgroup of the MIMIC dataset for the IHM task. Statistics of **(b)** prediction class distribution, **(c)** racial group distribution, and **(d)** age group distribution for the MIMIC IHM dataset. The MIMIC IHM training set consists of 45.1% female samples and 54.8% male samples. **(e)** Percentage of the minority class C1, Recall C0, and Recall C1 of each subgroup of the SEER dataset for the BCS task. Statistics of **(f)** prediction class distribution, **(g)** racial group distribution, and **(h)** age group distribution for the SEER BCS dataset. The SEER BCS training set consists of 99.4% female samples and 0.6% male samples.

To compute standard deviations, we repeat the machine learning training process multiple times, each time producing a machine learning model. Specifically, for BCS and LCS prediction tasks, we repeat the experiments five times. For the in-hospital mortality task, we repeat the experiments three times. Under these settings, average values and standard deviations are computed for all results except SHAP. Tables only show average results without error bars. All SHAP feature importance results (in the Supplementary Section) are based on the performance of a randomly selected machine learning model. For the decompensation prediction task, due to its high time complexity, we run the experiments once.

## Results

### Accuracy of majority and minority prediction classes without any bias correction

Without any bias correction, the original machine learning model demonstrates drastically different prediction capabilities for the majority prediction class C0 and the minority prediction class C1. Figure 2a shows recall values for both classes for various patient groups for in-hospital mortality (IHM) prediction and Figure 2e for predicting 5-year breast cancer survivability (BCS). For IHM, the recall value (0.61) for the minority class C1 is much lower than the recall of the majority class (0.88). For BCS, the recall C1 (0.67) is much lower than the recall C0 (0.93). This trend is consistently observed for various demographic groups, with a few exceptions of senior patients for BCS prediction. We further show detailed IHM predictions with the MIMIC III dataset for various subpopulations under 12 metrics in a heatmap in Figure 3a. 12% of non-death cases (class C0) in IHM prediction are wrong, whereas the missed mortality prediction (class C1) rate is much higher at 39%. For Black patients, while recall, precision, F1, and AUC-PR are all above or equal to 0.89 for class C0, the recall of class C1 is only 0.50, i.e., for every 100 Black patients who die in hospital, the model would mispredict 50 of them. A similar trend is observed for the BCS prediction results (Figure 3b). For the [40, 50) age group, the recall, precision, F1, and AUC-PR for majority prediction class C0 are all over 0.9, while for C1 merely 0.58, 0.48, 0.52, and 0.55 are observed, respectively.

**Figure 3:**
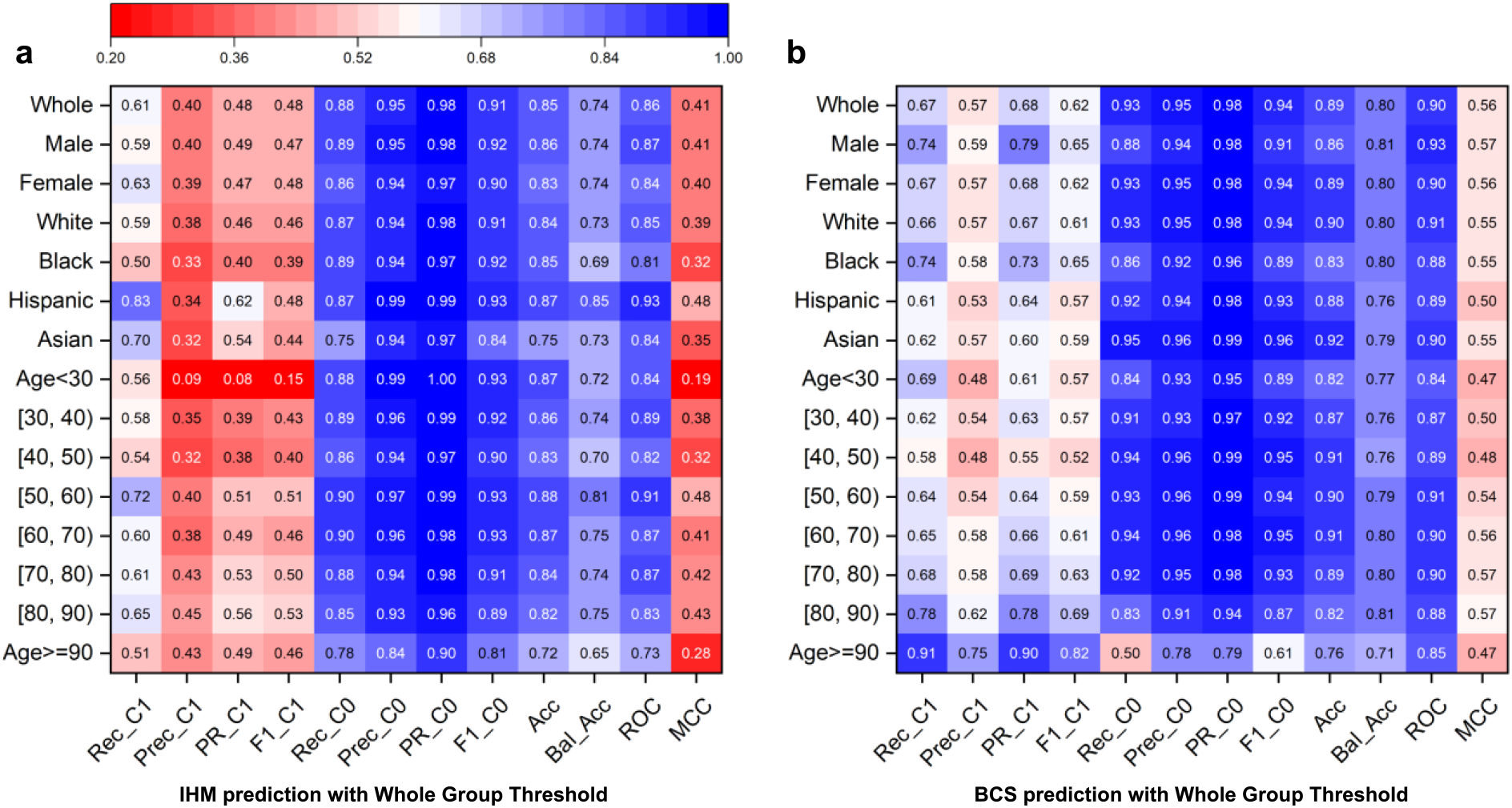
Prediction results under the original machine learning models (no bias correction) using one optimized threshold for all demographic groups. Rec_C1, Prec_C1, PR_C1, F1_C1, Rec_C0, Prec_C0, PR_C0, F1_C0, Acc, Bal_Acc, ROC, MCC stand for Recall Class 1, Precision Class 1, Area Under the Precision-Recall Curve Class 1, F1 score Class 1, Recall Class 0, Precision Class 0, Area Under the Precision-Recall Curve Class 0, F1 score Class 0, Accuracy, Balanced Accuracy, Area under the ROC Curve, Matthews Correlation Coefficient (MCC), respectively. **(a)** Prediction results for the IHM prediction. Class 1, representing death after staying 48 hours in intensive care units at the hospital, is the minority prediction class. Class 0, representing survival after staying 48 hours in intensive care units, is the majority prediction class. **(b)** Prediction results for the BCS prediction. Class 1, representing death 5 years after a breast cancer diagnosis, is the minority prediction class. Class 0, representing survival after 5 years, is the majority prediction class.

### Accuracy across demographic subgroups without bias correction

The original model also shows different prediction capabilities for specific demographic groups. For the IHM prediction (Figure 3a), Black patients have the lowest minority class C1 recall (0.50), lower than the whole group (0.61) and Hispanic patients (0.83). The difference among C1 recalls of various age groups is relatively smaller, all values in the range of [0.51, 0.72]. Most subgroups have somewhat similar C1 precision values, except the age <30 group. Young patients under 30 have a very low C1 precision of 0.09 in the IHM prediction, substantially lower than the whole population (0.40). This prediction deficiency is also reflected in the MCC metric, which is 0.19 for the age <30 group (Figure 3a). For the BCS task (Figure 3b), the minority class C1 recall (0.58) of age group [40,50) is only 64% of that of the 90+ age group (0.91), resulting in a large 0.33 difference. [40,50) and <30 groups have the lowest C1 precision; the 90+ age group has the highest. For the BCS prediction, accuracy difference across different racial groups also exists but appears less pronounced. The largest C1 recall difference is 0.13 between Hispanic (0.61) and Black (0.74). C1 precisions are all in the range of [0.53, 0.58].

Both gender groups perform similarly in both tasks, even though male patients only account for 0.6% of the samples in the SEER dataset for BCS prediction. Young patients under 30 account for only 0.6% and 4% in SEER (Figures 2h) and MIMIC III datasets (Figure 2d), respectively. Their predictions are consistently poor. Despite the large difference in minority class C1 performance, majority class C0 precisions and recalls are consistently high for all subgroups, with most values above 0.85. Despite small sample sizes, some demographic groups (e.g., 90+ groups in BCS prediction) have high prediction accuracies even without sampling.

### Metrics for imbalanced data

For imbalanced datasets, commonly used metrics such as AUC-ROC and accuracy are deceptive and do not reflect minority class performance. These metrics may show misleadingly higher values, even when the performance of the minority class is poor. The overall accuracy and AUC-ROC values are consistently high (> 0.80 in most cases, Figure 4) across different subgroups, even when minority class C1’s performance is less optimistic. None of the MCC values in Figure 3a exceeds 0.5 and the F1-score is only 0.39 for Black patients in IHM prediction.

**Figure 4:**
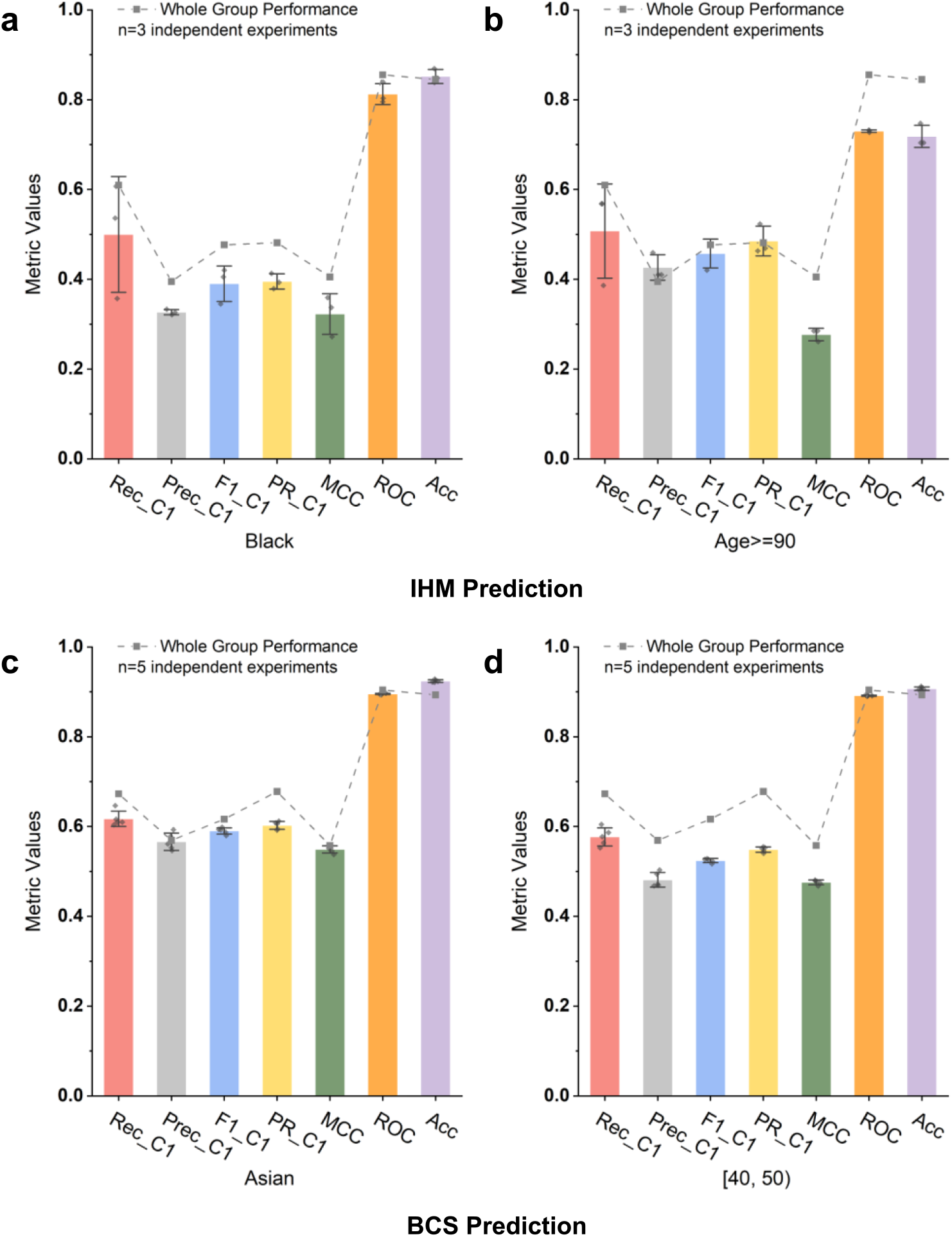
Comparison of whole-population metrics with minority-class-specific metrics. Some whole-population metrics (e.g., AUC ROC and accuracy) are misleading for the minority class. These deceptive metrics show high values, whereas the prediction is weak for the minority class. **(a)** Black subgroup performance for IHM prediction. **(b)** Age>=90 subgroup performance for IHM prediction. **(c)** Asian subgroup performance for BCS prediction. **(d)** Age [40,50) subgroup performance for BCS prediction.

Accuracy and AUC-ROC values are dominated by the overwhelmingly high precision and recall (> 0.85 in most cases) of the majority prediction class C0. Thus, these commonly used metrics in prediction do not reflect the minority class performance under data imbalance. In biased datasets, AUC-ROC is no longer sufficient, as it covers both classes with one dominating class. This deficiency is well established in the machine learning literature^42–44^, where multiple previous studies pointed out that AUC-ROC gives an overly optimistic view of imbalanced classification. Our work points out the severity of the metrics issue in digital health applications. In contrast to the overly optimistic AUC-ROC and accuracy metrics, MCC is a more sensitive metric and reflects prediction deficiencies in this type of imbalanced setting. By definition, MCC values range from [-1, 1], with 0 indicating the performance of a random classifier. Metrics reporting an individual class are also necessary to include.

### DP reduces accuracy disparity among demographic subgroups

We use relative disparity (defined in Equation 1) as a metric to quantify performance gaps across demographic subgroups under various machine learning conditions, including the original model (without any bias correction), DP bias correction, and existing sampling methods. Relative disparity measurement below 1.25 is considered fair, following the 80% rule for assessing disparate impact^45^. Our results show that machine learning models trained with our DP bias correction method exhibit the smallest racial and age disparities in most cases (Figure 5). For balanced accuracy and C1 recall of both IHM and BCS tasks, most of DP’s relative disparity values are in the fair range (1.25 and lower), substantially reducing the gap in the original model. Specifically, DP has a 14.8% to 23.9% improvement over the original model in terms of the relative disparity of C1 recall. DP method also reduces MCC disparity the most for in-hospital mortality prediction (Figure 5c).

**Figure 5:**
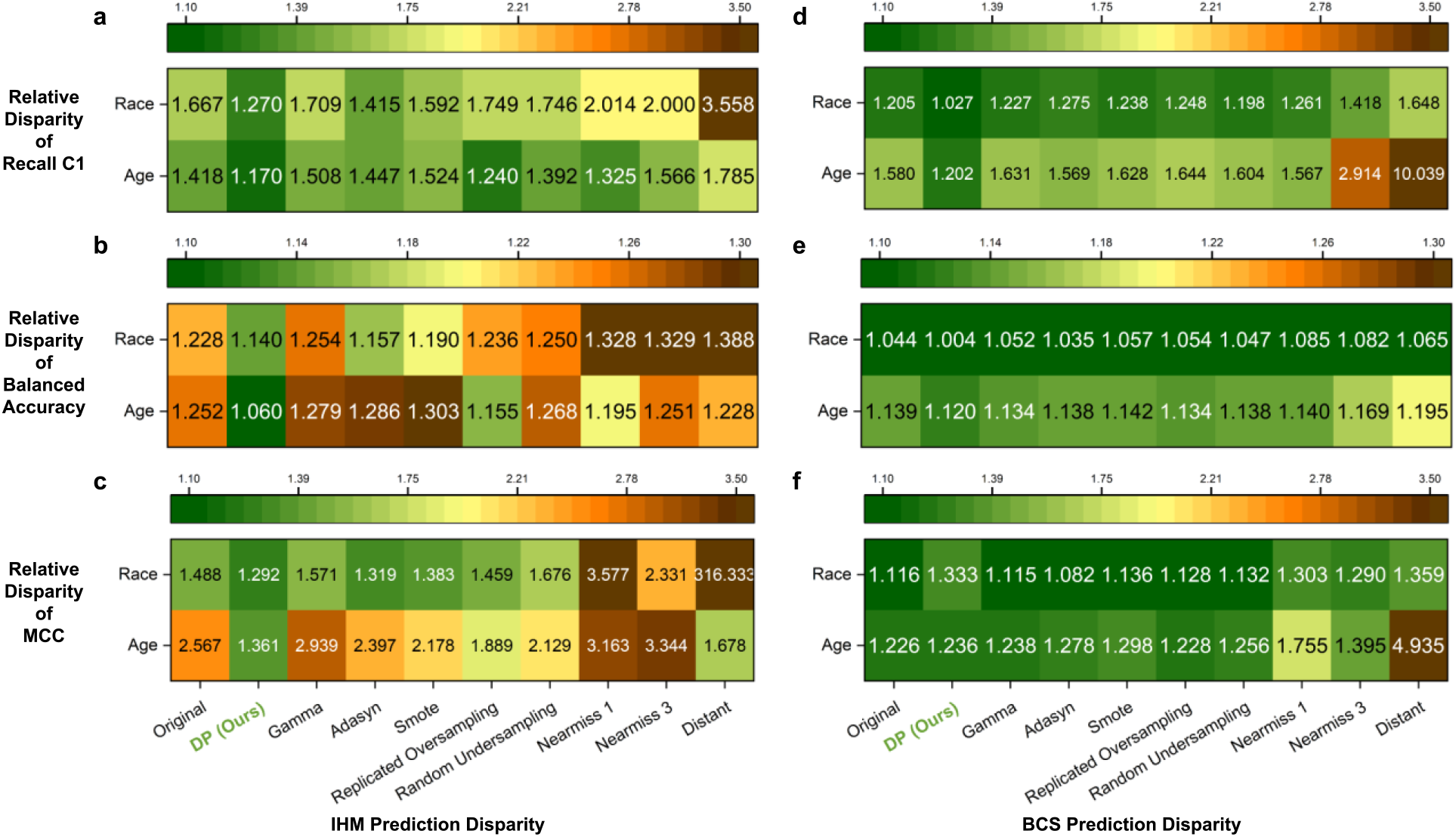
Relative disparity among racial and age groups under various sampling conditions, including DP and the original machine learning model without any sampling. Relative disparity of MIMIC III IHM prediction in terms of **(a)** minority class recall, **(b)** balanced accuracy, and **(c)** Matthews correlation coefficient (MCC). Relative disparity of SEER BCS prediction in terms of **(d)** minority class recall, **(e)** balanced accuracy, and **(f)** Matthews Correlation Coefficient (MCC). DP performs the best in reducing the relative disparity across subgroups (i.e., showing the lowest disparity values) compared to the original model and models with other existing sampling methods for both tasks.

In contrast, all three state-of-the-art sampling methods (namely, Gamma, Adasyn, and SMOTE) fail to substantially reduce the racial and age disparities in the IHM task, with some models (e.g., Gamma) exacerbating disparity. Undersampling methods (especially Distant) perform worse than oversampling methods. When compared to the eight existing methods, DP reduces racial disparity by 10.2% (ADASYN) to 64.3% (Distant) and age disparity by 5.6% (Replicated Oversampling) to 34.5% (Distant), in terms of the minority C1 recall for IHM prediction (Figure 5a). Balanced accuracy (Figure 5b) and MCC results (Figure 5c) follow a similar trend. The Distant method’s MCC race disparity is high (316.3), due to its extremely low MCC score for Hispanic patients (0.001).

While the racial and age disparity is less severe for BCS prediction, the advantage of DP can still be observed. Overall, DP shows 14.3% (random undersampling) to 37.7% (Distant) improvement among racial groups and 23.3% (NearMiss 1) to 88.0% (Distant) improvement for age groups in terms of C1 recall (Figure 5d) compared to existing sampling methods.

### Mitigation solely based on adjusting thresholds

We also test whether or not threshold tuning alone can boost the performance of demographic subgroups and reduce disparity. Specifically, we compare the prediction performance under the whole group threshold and subgroup thresholds, which are described in the *Methods* section. Prediction results under the original machine learning models (no bias correction) using different optimized thresholds for different demographic groups are shown in Supplementary Figure 2. For the IHM task, the performance differences between using the whole-group threshold and subgroup threshold are small (< 0.1), in terms of C1 precision and recall, for subgroups with relatively large sizes (e.g. middle-aged patients). However, for other smaller subgroups (e.g. young patients with age<30), the performance decreases. A likely reason is overfitting, i.e., the threshold selected based on a small sample size in the validation set is not optimal on the test set, due to the small sample sizes. BCS results follow similar patterns. Thus, threshold adjustment alone is insufficient for the data imbalance and accuracy disparity problems.

### Subpopulation-based vs. whole-population-based sampling

Existing sampling solutions do not differentiate subpopulations. We found such whole-population-based sampling methods decrease the performance of some underrepresented groups. We compare DP with two common sampling techniques (i.e., random undersampling and SMOTE) with four demographic groups (namely, Black, Asian, age < 30, 90+ for the IHM task and Hispanic, Asian, age <30, 90+ for the BCS task). These groups are chosen because of their low performances under the original machine learning model. DP consistently boosts the performance of most underrepresented demographic groups (Figure 6). For IHM, DP improves the original model’s C1 recall by 6.0% to 38.0%. This improvement is up to 29.6% for BCS. In contrast, this consistent improvement is not observed in the other two methods. For example, for the IHM task, although the undersampling technique boosts the balanced accuracy for Asian patients, the performances of Black and age 90+ subgroups slightly decrease (Figure 6b). For the BCS task, SMOTE slightly decreases the C1 recall for the Hispanic, Asian, and age [40,50) groups (Figure 6c). We note that for the age <30 subgroup, DP’s balanced accuracy drops (Figure 6d), which is due to a decrease in the majority class C0 recall. The complete comparison results with the 8 existing sampling methods and the original machine learning model without any sampling are shown in Supplementary Figure 3.

**Figure 6:**
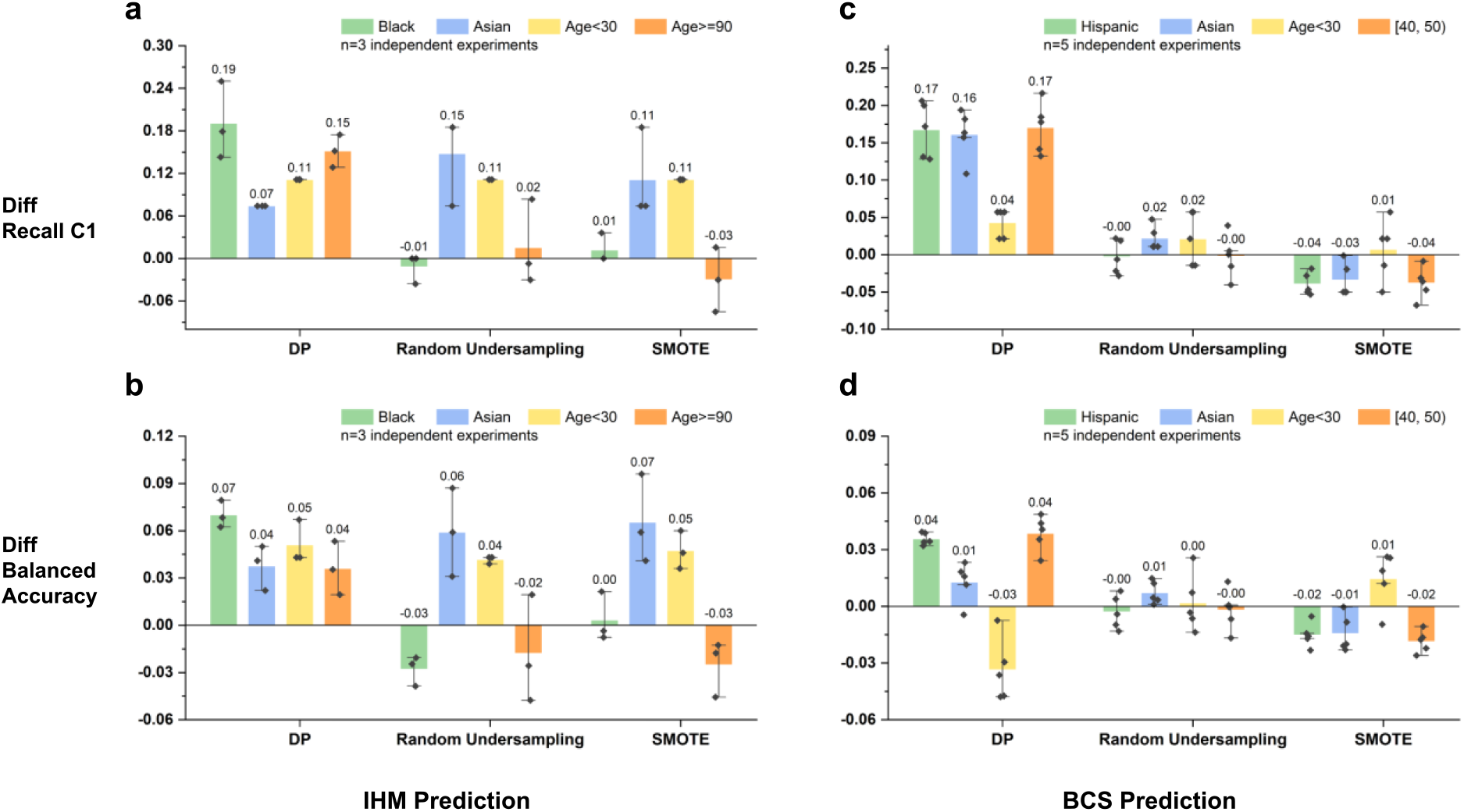
Performance comparison of DP and two representative sampling techniques (namely, random undersampling and SMOTE) over the original model for four demographic subgroups with poor original performance. Positive values indicate performance improvement, and negative values indicate performance degradation from the original model. The error bars represent the standard error of the experiment results. Performance comparison **(a)** in terms of recall C1 for IHM prediction with the MIMIC III dataset, **(b)** in terms of balanced accuracy for IHM prediction with the MIMIC III dataset, **(c)** in terms of recall C1 for the BCS prediction with the SEER dataset, **(d)** in terms of balanced accuracy for the BCS prediction with the SEER dataset.

For subgroups with lower original performance, DP brings stronger C1 recall improvements. We show this trend in Figure 7, where we compare the minority class recall between the original model with subgroup threshold and the DP model trained for each subgroup. For the IHM task, DP improves the C1 recall by 200.4%, 163.4%, and 75.2%, respectively, for the age <30, Black, and Asian patients (Figure 7a). Similarly, for the BCS task, C1 recall of DP is 30.7%, 27.3%, and 27.1% higher than the original model with subgroup threshold for age [40, 50), Hispanic, and Asian patients, respectively (Figure 7b).

**Figure 7:**
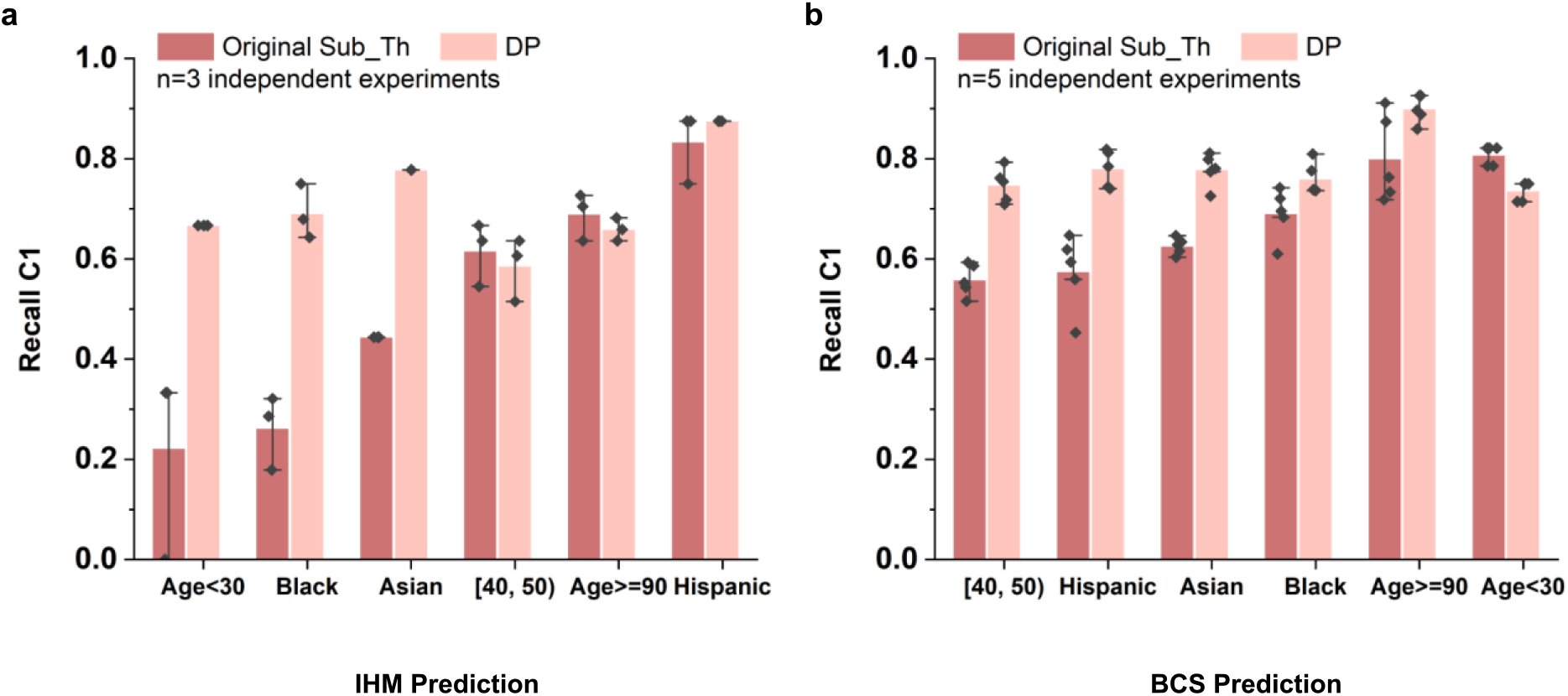
Performance of DP and subgroup-threshold-based original model in terms of minority class recall for in-hospital mortality (IHM) prediction and 5-year breast cancer survivability (BCS) prediction. Darker red color represents the original model performance using subgroup optimized threshold and lighter red color represents DP performance. The error bars represent the standard error of the experiment results. DP’s improvements are stronger when the original recall C1 values are relatively low, partly because DP selects machine learning models based on balanced accuracy. Model performance comparison for **(a)** IHM prediction task and **(b)** BCS prediction task of 6 different racial or age subgroups. For the IHM prediction task, the standard deviation values for DP are between 0 and 0.051. For the BCS prediction task, the standard deviation values for DP are between 0.017 and 0.033.

### Impact of specialized machine learning models on prediction outcomes

In our cross-group experiments, we use the DP model trained for demographic group A (e.g., Black) to predict group B (e.g., Hispanic). The aim is to evaluate the impact of different machine learning models on prediction outcomes. We perform both cross-race and cross-age-group experiments for BCS prediction (Figure 8) and IHM prediction (Supplementary Figure 4), which involve 3 underrepresented races and 3 underrepresented age groups. For 5 out of the 6 DP models in BCS prediction, the minority class C1 recall is the highest when the matching DP model is applied, i.e., when the race or age group of patients being predicted matches the race or age group that the DP model is trained for. For example, when predicting Asian patients’ breast cancer survivability, the DP Asian model (0.78) outperforms the DP Black model (0.55), DP Hispanic model (0.58), and the original model without DP (0.62), in terms of minority class C1 recall (Figure 8a). Similarly, the balanced accuracy is the highest when DP Asian model is applied to predict Asian patients (Figure 8c). In the cross-age-group experiment, this trend is also observed. For example, DP [40, 50) model substantially outperforms the other three models when predicting patients in the [40, 50) age range. Its recall C1 is 0.75, whereas the DP <30, DP 90+, and the original models give 0.56, 0.52, and 0.58, respectively (Figure 8b). However, the DP 90+ model does not show an advantage, as the original model gives a slightly higher recall C1 and the DP [40, 50) gives the highest balanced accuracy when being applied to 90+ patients.

**Figure 8:**
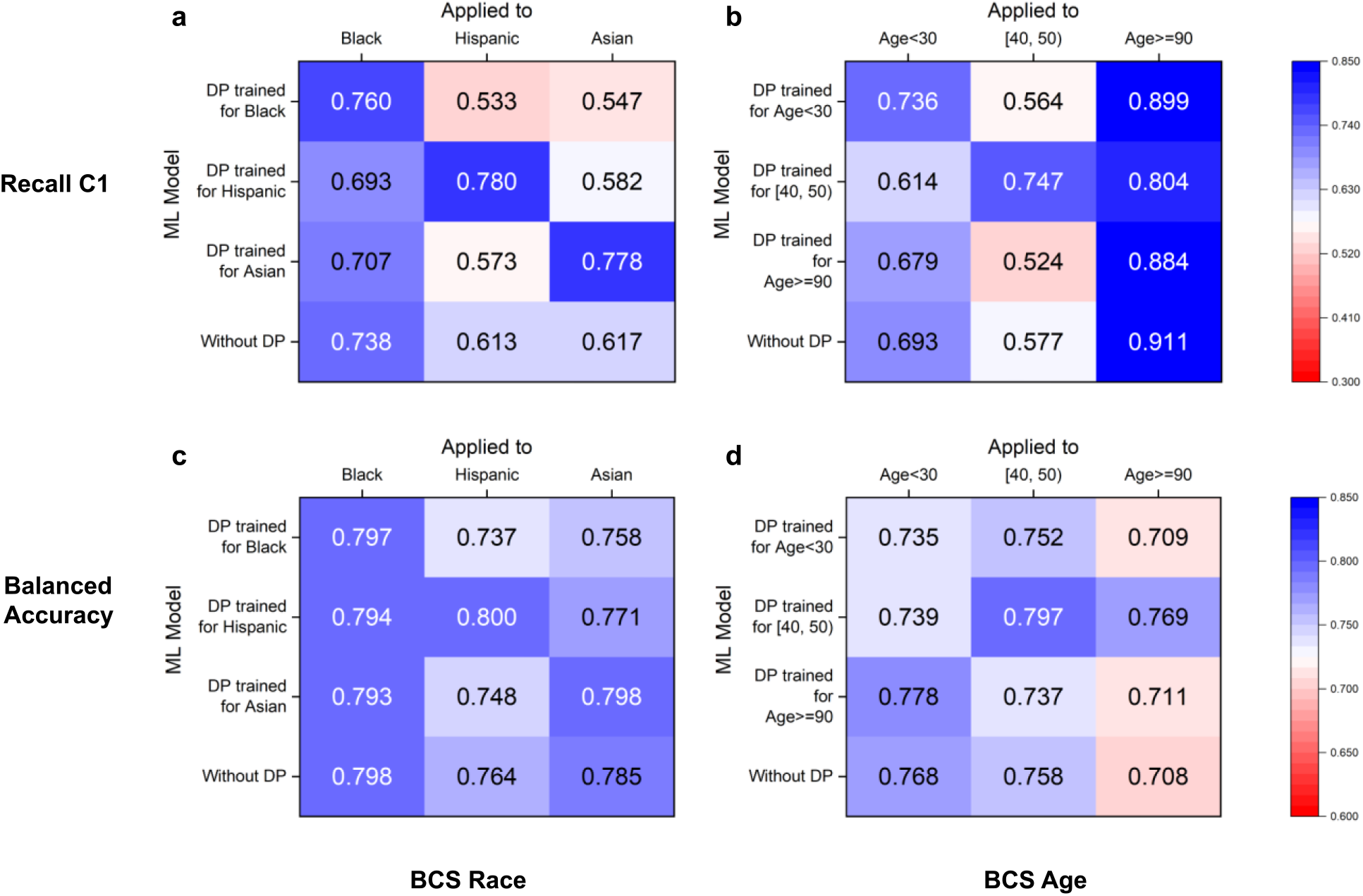
DP’s cross-group performance under various race and age settings for recall C1 and balanced accuracy in BCS prediction. In subfigures, each row corresponds to a DP model trained for a specific subgroup. Each column represents a subgroup that a model is evaluated on. The values on the diagonal are the performance of a matching DP model, i.e., a DP model applied to the subgroup that it is designed for. The last rows show the group’s performance in the original model. To prevent overfitting, our method chooses optimal thresholds based on whole group performance. DP cross-group performance for **(a)** race subgroups and **(b)** age subgroups for the BCS prediction in terms of recall C1. DP cross-group performance for **(c)** race subgroups and **(d)** age subgroups for the BCS prediction in terms of balanced accuracy.

For IHM prediction, DP models’ advantage is observed in 3 out of the 6 groups (for Black, <30, and 90+ groups), which is less pronounced than BCS prediction (Supplementary Figure 4). In the cross-age-group experiment, both DP <30 and 90+ models demonstrate advantages. For Hispanic and Asian patients, the DP Black model gives the best recall C1, higher than DP Hispanic and DP Asian models.

### Decompensation prediction and 5-year lung cancer survivability (LCS) prediction

We repeat the experiments for the other two tasks, decompensation prediction, and 5-year lung cancer survivability (LCS) prediction, and observe similar patterns. For decompensation prediction on the MIMIC III dataset, the minority class C1 represents patients whose health condition deteriorates after 24 hours. Without any bias correction, C1 recall is merely 0.40 and 0.39 for Black (Supplementary Figure 5a) and age 90+ patients (Supplementary Figure 5b), respectively, while C0 recalls are near perfect (Supplementary Figure 6a). Prediction accuracy also differs across demographic subgroups, e.g., C1 precision is 0.46 for age 90+ patients and 0.13 for age <30 patients (Supplementary Figure 6a). For LCS prediction on the SEER dataset, the minority Class 1 represents patients who survive lung cancer for at least 5 years after the diagnosis. Without any bias correction, the recall, precision, and AUC-PR are all above 0.93 for Class 0, while the values for Class 1 are lower at 0.65, 0.61, and 0.67, respectively, for Black patients (Supplementary Figure 5c). Regarding each demographic subgroup, the original model catches all survival cases (minority class in LCS) in the age <30 group, however, it misses 40% and 70% of the survival cases in age [80, 90) and 90+ groups, respectively (Supplementary Figure 6b). Results on subgroup thresholds (Supplementary Figure 7) follow a trend similar to the earlier IHM and BCS findings.

Sampling results for the decompensation and LCS prediction tasks are shown in Supplementary Figures 8-11. For decompensation prediction, we apply the two most commonly used sampling techniques, random undersampling (RUS) and replicated oversampling (ROS). We have to exclude other sampling techniques as their pairwise quadratic distance computation is expensive for 2,377,768 patients’ time series training dataset. After applying DP bias correction, the minority class C1 recall for most subgroups consistently improves (Supplementary Figures 8a and 9a). The DP improvements are higher than applying RUS and ROS (Supplementary Figures 8a and 8b). Regarding fairness for the decompensation task, the relative disparity of DP is lower than or comparable to other sampling approaches for most cases (Supplementary Figure 10a-c), which is consistent with the trend observed in Figure 5. We examine an exceptional case for race groups in terms of recall, where the high Hispanic group performance (0.76) increases the disparity value (Supplementary Figure 10a). For the LCS prediction task, the results of applying DP and other sampling methods follow a similar pattern as the BCS prediction. For sampling’s fairness comparison, NearMiss1 undersampling shows the lowest relative disparity for age groups in terms of C1 recall (Supplementary Figure 10d). While NearMiss1 brings C1 recall of all age groups to a relatively good range of [0.63, 1.00], its C1 precision ([0.03, 0.54]) is poor. NearMiss1’s MCC age disparity shown in Supplementary Figure 10f is high (5.36), as MCC is a more comprehensive and sensitive metric. Additional sampling comparisons can be found in Supplementary Figure 11.

We also conduct cross-group experiments for the LCS task and the decompensation task. For the LCS prediction, 4 out of 6 matching DP models (i.e., Black, Hispanic, Asian, and age [80, 90) groups) show an advantage in terms of both C1 recall and balanced accuracy (Supplementary Figure 12). Two exceptions are the age [30, 40) and 90+ groups. The original model performs the best for the age [30, 40) subgroup; the [80, 90) DP model outperforms others on the age 90+ patients. Supplementary Figure 13 shows that matching DP models show some degree of advantage in 4 out of 6 settings for the decompensation task.

### Reweighting and feature importance

The standard reweighting models, where reweighting does not differentiate subpopulations, perform almost identically to the original model when applied to Asian and age [40, 50) patient groups (Supplementary Figure 14). This performance similarity between the standard reweighting model and the original model is also observed in LCS prediction for Black and 90+ patient groups (Supplementary Table 2). In contrast, prioritized reweighting, where new weights are optimally placed on a specific group of patients, boosts C1 recall in BCS prediction for Asian patients from 0.617 to 0.802 (Supplementary Figure 14a) and from 0.577 to 0.763 for age [40, 50) patients (Supplementary Figure 14b). This boost is comparable to DP’s performance. DP and prioritized reweighting also exhibit comparable performances under other metrics (Supplementary Figure 14).

Cancer survivability prediction on the SEER dataset includes age and race features. Under SHAP-avg, age-related features rank at the very top for all BCS and LCS prediction models (Supplementary Figures 15 and 16). Race-specific DP and prioritized reweighting models rank race features higher than the original and the standard reweight models in the BCS prediction. For example, race recode A and Y are the top 5^th^ and 6^th^ features in both the DP Asian model and the prioritized reweighting model for Asians (Supplementary Figures 15b and 15e). For LCS prediction, the race feature ranks 16^th^ in the DP Black model (Supplementary Figure 16c). In contrast, race is not among the top 18 features for the original or standard reweight models. For BCS and LCS tasks under SHAP-avg, top clinical features include the number of positive lymph nodes examined, tumor size and site, grade, and stage (Supplementary Figures 15 and 16), which are expected.

Race-specific models and age-specific models show different top features or have different orderings of top features for BCS and LCS predictions. For example, [40, 50)-specific models (Supplementary Figures 15c and 15f) have multiple age-related top features, but do not have race features in the top ranks. For DP and prioritized reweighting models, their top features for the same demographic group appear very similar, which is consistent with their similar prediction performance. For example, for BCS prediction, DP and prioritized reweighting models for Asian have identical top 8 features; the models for the [40, 50] age group have identical top 7 features (Supplementary Figure 15).

For IHM and Decomp tasks under SHAP-avg, the top features of the DP models and the original models are similar, slightly differing in their feature ordering (Supplementary Figures 17 and 18). For example, for IHM prediction DP age 90+ model ranks weight at the 4^th^ position, slightly higher than its ranking in the DP Black and the original models (both at the 7^th^ position). This observation may suggest that overweight in older patients is more likely to cause serious consequences. Following the existing benchmark^17^, our IHM and decompensation predictions only use 17 clinical features and exclude race and age information in MIMIC III. We found that SHAP-sum identifies very different top features from SHAP-avg, highlighting categorical features due to their multiple one-hot encoding representations for machine learning. We show the SHAP-sum feature ranking of IHM prediction in Supplementary Figure 19. We discuss them in the next section.

For BCS and LCS predictions, default MLP model setting gives performances comparable to the other two neural network structures, in terms of prediction accuracy (Supplementary Table 4) and relative disparity (Supplementary Table 5).

## Discussion

Our findings empirically demonstrate multiple deficiencies of typical machine learning prognosis procedures when they are applied to imbalanced medical datasets. One deficiency is that the weak performance of underrepresented patients may be eclipsed by the whole population performance and not accurately reported. Underrepresentation is two-fold: *i)* demographic subgroups and *ii)* the minority prediction class. The low accuracy problem is particularly severe when a patient belongs to both categories. For example, for the IHM prediction, Black patients’ C1 recall (0.50) is 18% lower than the whole group (0.61) (Figure 3). Low recalls in the disease group can lead to underestimation of risks, missed treatment opportunities, or life-threatening wrong prognoses. In addition, racial and age disparities in machine-learning-based prognoses are also observed. Conceptually, these findings are consistent with what other AI fairness studies have reported, e.g., for face recognition^28, 39^. Thus, besides conventional machine learning accuracy metrics, fine-grained single-class metrics and fairness metrics need to be used, which will provide important insights into how well machine learning models respond to different types of patients.

Our work also reveals that the machine learning model computed based on the whole population may not be the optimal model for an underrepresented demographic subgroup. Conventional machine learning prognoses follow a one-model-predicts-all-demographics paradigm. Similarly, all existing sampling methods are also designed to oversample or undersample across all demographics. Our results show that the existing one-model-for-all-demographics approaches including sampling methods are not well equipped to achieve good fairness performance when the training data has biases.

A key contribution of our work is to systematically compare the conventional one-model-fits-all approach with a new double-prioritized (DP) bias correction approach, where specialized prognosis models are trained for minority prediction class patients of a certain race or age. Conceivably, it is challenging to train a single machine learning model that optimizes for all demographic groups. In contrast, the DP bias correction technique allows one to train models for specific demographic groups, not having to use the same model for the entire patient population. The key enabler of DP is demographic-specific sampling, i.e., selectively enriching the number of samples in the minority prediction class (C1). Training a specific machine learning model for some patient groups is necessary. For example, the oldest-old age group (typically defined as 85+)^46^ is a growing population in the US^47^. However, our study shows that 90+ patients’ recall C1 value (0.51) in the mortality prediction is 16% lower than the whole group (0.61) in the original model. Prioritized bias correction is highly effective for improving C1 recalls of demographic subgroups who are underrepresented in the training data, e.g., DP’s recall C1 is 0.66 (29.4% improvement) for 90+ patients in mortality prediction.

Our cross-race and cross-age-group experiments evaluate the impact of specialized machine learning models on prognosis accuracy. Overall, 16 out of the 24 (67%) matching DP models across the four tasks demonstrate an advantage over non-matching models, where the matching DP models (i.e., sample enrichment matches the test group’s demographics) achieve the highest recall C1 performance. Out of the 16 DP models, 8 of them are race models and 8 of them are age models (Supplementary Table 3). These findings confirm that algorithms matter in prognosis prediction and different model choices can significantly impact accuracy. These results also indicate the need for training specialized machine learning models for underrepresented patient groups.

Model specialization still needs to rely on the whole group samples. Training a model solely based on particular subgroup samples (e.g., Black patients) gives poor results, worse than the original model on almost all metrics, due to small sample sizes. This result (not shown) suggests the importance of involving all samples in the training, which forms a necessary starting point for further model optimization. The whole population training takes full advantage of shared features before subsequent model specialization. We also compare the original machine learning model under two calibration conditions for the IHM prediction -- calibration based on the whole group or calibration based on a specific subgroup. The results are similar for most cases (Supplementary Figure 20). For several underrepresented groups (e.g., Black, Asian, age <30, and age [30,40)), their recalls are lower if we apply subgroup calibration. Thus, our experiments are conducted under the whole group calibration condition, unless otherwise specified. Similarly, when applying subgroup optimized thresholds, we observe small performance changes for relatively large subgroups and decreased performance for the smaller ones (Supplementary Figures 2 and 7). One possible reason is that the selected threshold is overfitted to the small sample size in the validation set, resulting in lower testing performance. Therefore, we use a whole-group-based threshold on our DP and other bias correction experiments.

Prioritized reweighting results further confirm the need for designing subpopulation-specific bias correction mechanisms in machine learning. The prioritized reweighting method described in this paper is new. It puts more weights on a subset of C1 samples, as opposed to applying the same weight to all C1 samples. Prioritized reweighting performs similarly to the DP method (Supplementary Figure 14). This similarity is expected for two reasons. First, the workflow of the prioritized reweighting method is designed to mimic DP to emphasize specific subgroups’ C1 samples. Their only difference is that the former increases the weights and the latter adds more samples. Second, literature shows that reweighting and sampling approaches are statistically equivalent if operating under similar conditions^38^. In contrast, the standard reweighting method, which reweights the entire C1 population, has a weaker effect in boosting recall C1 for specific subpopulations (Supplementary Figure 14). The standard reweighting performs almost identically to the original model for Asian and age [40, 50) patients in the BCS prediction.

When computing feature importance, our results show that the SHAP-avg approach is more appropriate for one-hot encoded categorical features that have a large number of choices. When compared with SHAP-sum, SHAP-avg performs an additional step to normalize the importance value of a categorical feature. Without this step, categorical features with a large number of possible values are ranked high. For example, irrelevant features such as the patient’s state-county information and SEER registry (about data source) consistently rank high under SHAP-sum for the BCS and LCS predictions (results not shown), which is because of their hundreds of columns in the one-hot encoding representation. On the other hand, when the size of the category is small, the SHAP-sum ranking can be meaningful. For example, the Glasgow Coma Scale contains three categorical features, each with 4 to 6 options. SHAP-sum ranks the Glasgow Coma Scale (i.e., the extent of impaired consciousness) at the very top for all models in both the IHM prediction (Supplementary Figure 19) and decompensation prediction (not shown). Their ranks drop to the 7^th^ to 14^th^ positions under SHAP-avg. In this IHM case, both SHAP-sum and SHAP-avg methods give meaningful rankings. Further AI interpretability research will help develop a more systematic methodology for ranking one-hot encoded features.

DP bias correction does not boost the performance of the majority prediction class and may reduce the model’s overall performance if applied. In BCS prediction, death is the minority prediction class for most demographic groups. However, for the age 90+ group, nearly 60% of the patients died within 5 years, making death the majority prediction class in this subgroup. Thus, the original model’s C1 performance is good, in terms of recall (0.91), precision (0.75), AUC-PR (0.90), and F1 (0.82). In contrast, the class C0 performance for age 90+ is weaker, with 0.50 recall and 0.61 F1. Further increasing the number of C1 (death) cases would cause the data to be even more imbalanced. Thus, a key first step in DP is to identify the minority prediction class and the underrepresented demographic subgroups in the training dataset.

Our results show that DP can mitigate racial and age disparities introduced by data underrepresentation in training machine learning models, better than the existing 8 sampling methods being compared. However, data imbalance is only one source of disparity. For example, the diagnosis and treatment conditions may vary across different demographic subgroups and affect data quality. These variations may also contribute to the disparity observed across groups. Eliminating such more fundamental and systemic medical biases is beyond the scope of technical solutions.

In summary, because underrepresentation is prevalent in clinical medicine, our findings likely have broad implications beyond the specific datasets and demographic groups studied. Fully recognizing accuracy disparities associated with imbalanced data will help reduce life-threatening prediction mistakes. Vast accuracy gaps exist between minority C1 and majority C0 classes and across some demographic subgroups. When training and testing machine learning models, using multiple metrics is crucial, including balanced accuracy and separate metrics for the two prediction classes. Commonly used metrics, namely AUC-ROC and accuracy, are heavily influenced by the majority class and may fail to reflect the minority class performance when the dataset is imbalanced. DP bias correction is applicable to medical datasets, where data imbalance may be a source of accuracy disparity. The method is not designed to address non-representational disparities, e.g., underdiagnosis and measurement bias. Future directions include further enhancing the interpretability of machine learning prognosis models, as well as exploring how data underrepresentation impacts the quality of medical image analysis and mutation-based evolutionary computation^48^.

## Data Availability

The MIMIC III and SEER data used in this study are not publicly downloadable but can be requested at their original sites. Parties interested in data access should visit the MIMIC III website (https://mimic.physionet.org/gettingstarted/access/) and the SEER website (https://seer.cancer.gov/data/access.html) to submit access requests.

https://seer.cancer.gov/data/access.html

## Contributors

DY conceived and designed the study. DY and CL conceived the DP bias correction method. DY designed the cross-group model experiments. DY, SA, and WS designed the prioritized reweighting method. SA defined the relative disparity metric. SA conducted experiments on MIMIC III and analyzed the data. WS conducted experiments on SEER and analyzed the data. SA and WS cross-checked the validity of each other’s data. SA and WS designed the sampling comparisons. SA, WS, and DY wrote the manuscript. CL and CBN provided strategic guidance. All authors proofread the manuscript and provided feedback.

## Code Availability

We have released all our code used on GitHub. The directory contains the preprocessing code for training data generation for DP, as well as result processing regarding model selection and subgroup result extraction steps. https://github.com/ShaAfr/underrepresentation_in_clinical_dataset

## Supplementary Information

### Supplementary Equations

BCS Class 1: Patient does not survive more than 5 years after breast cancer diagnosis;

IHM Class 1: Based on the first 48 hours of ICU information, the patient dies in ICU

LCS Class 1: Patient survives more than 5 years after lung cancer diagnosis

Decomp Class 1: Patient’s health deteriorates after 24 hours

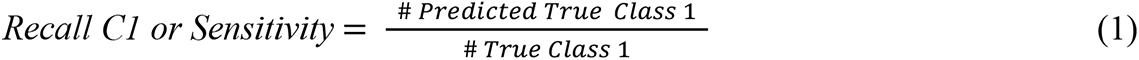

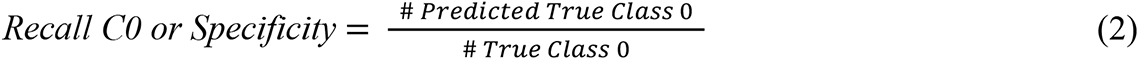

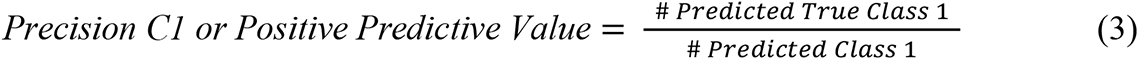

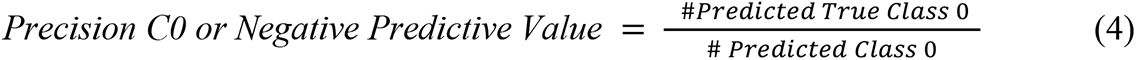

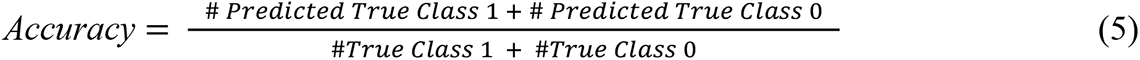

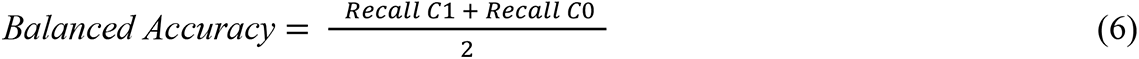

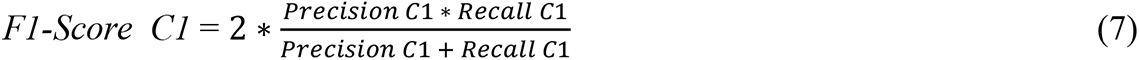

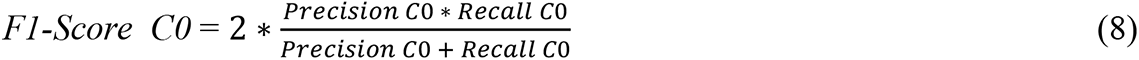

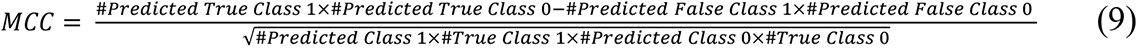

### Supplementary Figures and Tables

**Supplementary Figure 1:**
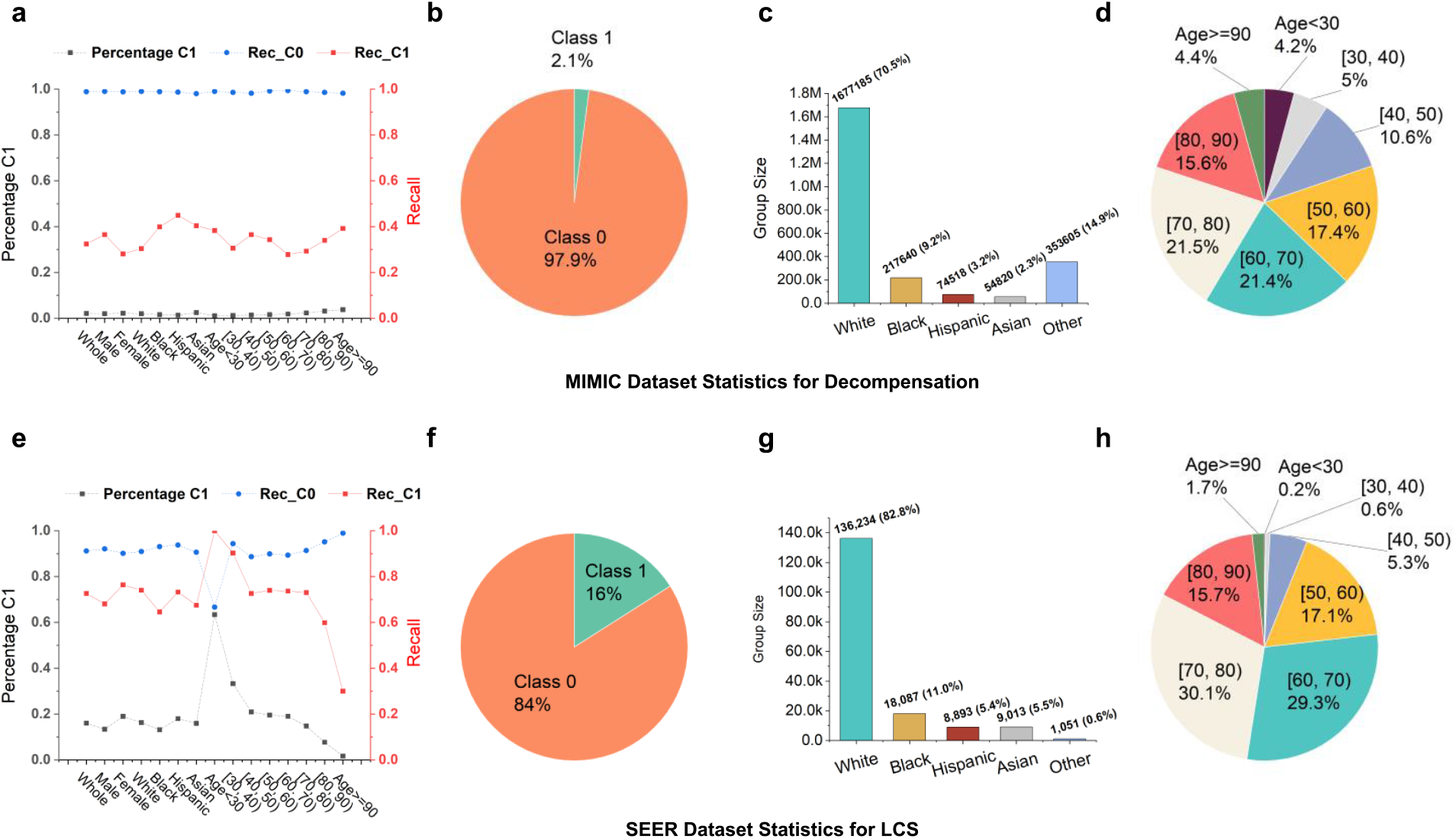
Recall values for both classes C0 and C1 and training data statistics for the decompensation and the 5-year lung cancer survivability (LCS) tasks. **(a)** Percentage of the minority class C1, Recall C0, and Recall C1 of each subgroup of the MIMIC dataset for the Decomp task. Statistics of **(b)** prediction class distribution, **(c)** racial group distribution, and **(d)** age group distribution for the MIMIC Decomp dataset. The MIMIC Decomp training set consists of 44.3% female samples and 55.7% male samples. **(e)** Percentage of the minority class C1, Recall C0, and Recall C1 of each subgroup of the SEER dataset for the LCS task. Statistics of **(f)** prediction class distribution, **(g)** racial group distribution, and **(h)** age group distribution for the SEER LCS dataset. The SEER LCS training set consists of 47.0% female samples and 53.0% male samples.

**Supplementary Figure 2:**
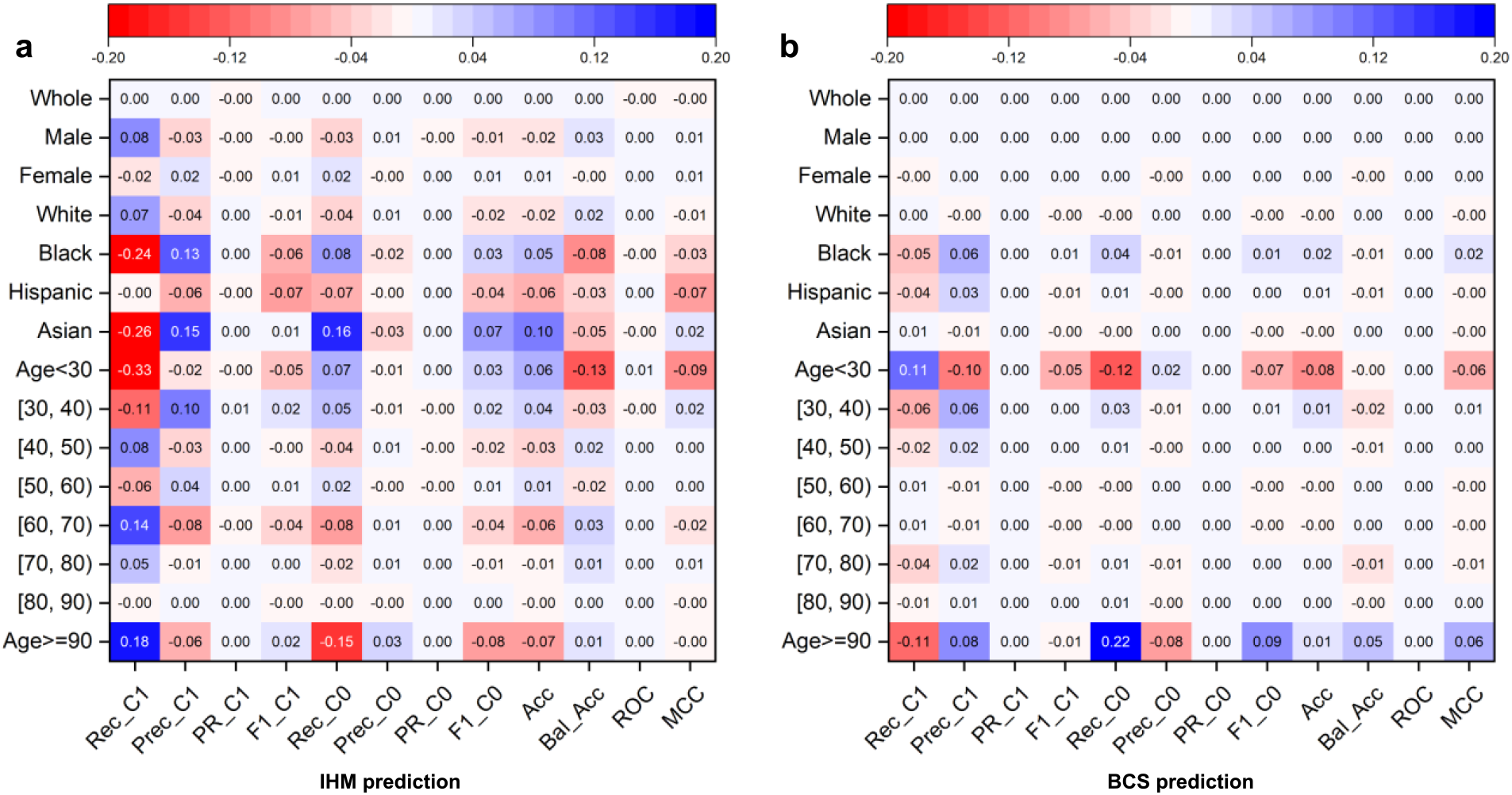
Differences in performance of the original machine learning models (no bias correction) using subgroup thresholds (i.e., different optimized thresholds for different demographic groups) and using the whole group threshold. Positive values mean that using a subgroup optimized threshold improves the performance. Rec_C1, Prec_C1, PR_C1, F1_C1, Rec_C0, Prec_C0, PR_C0, F1_C0, Acc, Bal_Acc, ROC, MCC stand for Recall Class 1, Precision Class 1, Area Under the Precision-Recall Curve Class 1, F1 score Class 1, Recall Class 0, Precision Class 0, Area Under the Precision-Recall Curve Class 0, F1 score Class 0, Accuracy, Balanced Accuracy, Area under the ROC Curve, Matthews Correlation Coefficient, respectively. The performance differences between the two settings are shown for **(a)** the IHM prediction and **(b)** the BCS prediction.

**Supplementary Figure 3:**
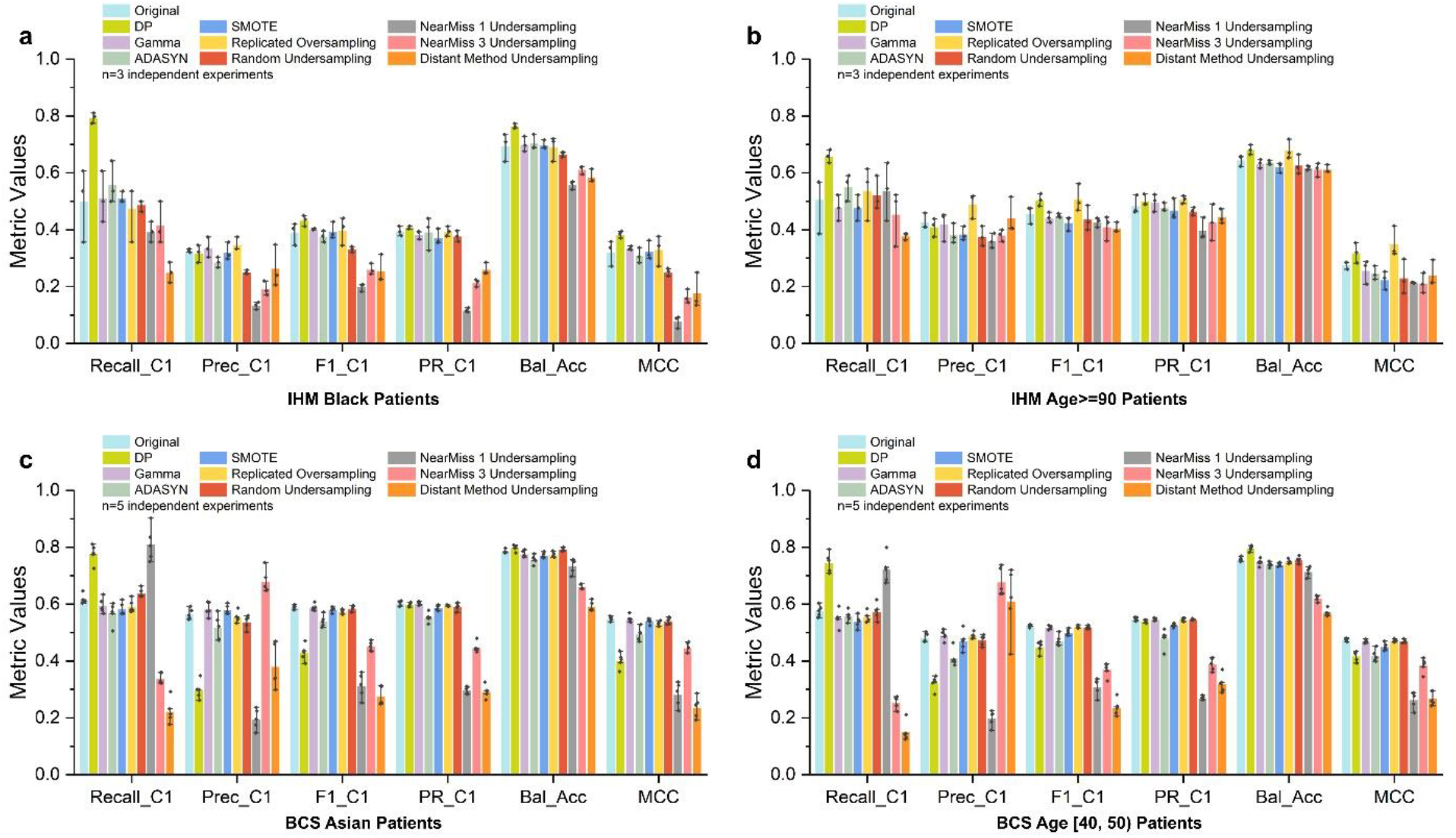
In-hospital mortality (IHM) prediction and 5-year breast cancer survivability (BCS) prediction under various sampling conditions, including DP and the original machine learning model without any sampling, in terms of minority class recall, precision, F1 score, AUC-PR, balanced accuracy, and Matthews Correlation Coefficient (MCC). Prediction results from the original model and different sampling models for **(a)** Black patients and **(b)** age>=90 patients in the IHM prediction with the MIMIC III dataset. Prediction results from the original model and different sampling models for **(c)** Asian patients and **(d)** age [40, 50) patients in the BCS prediction with the SEER dataset.

**Supplementary Figure 4:**
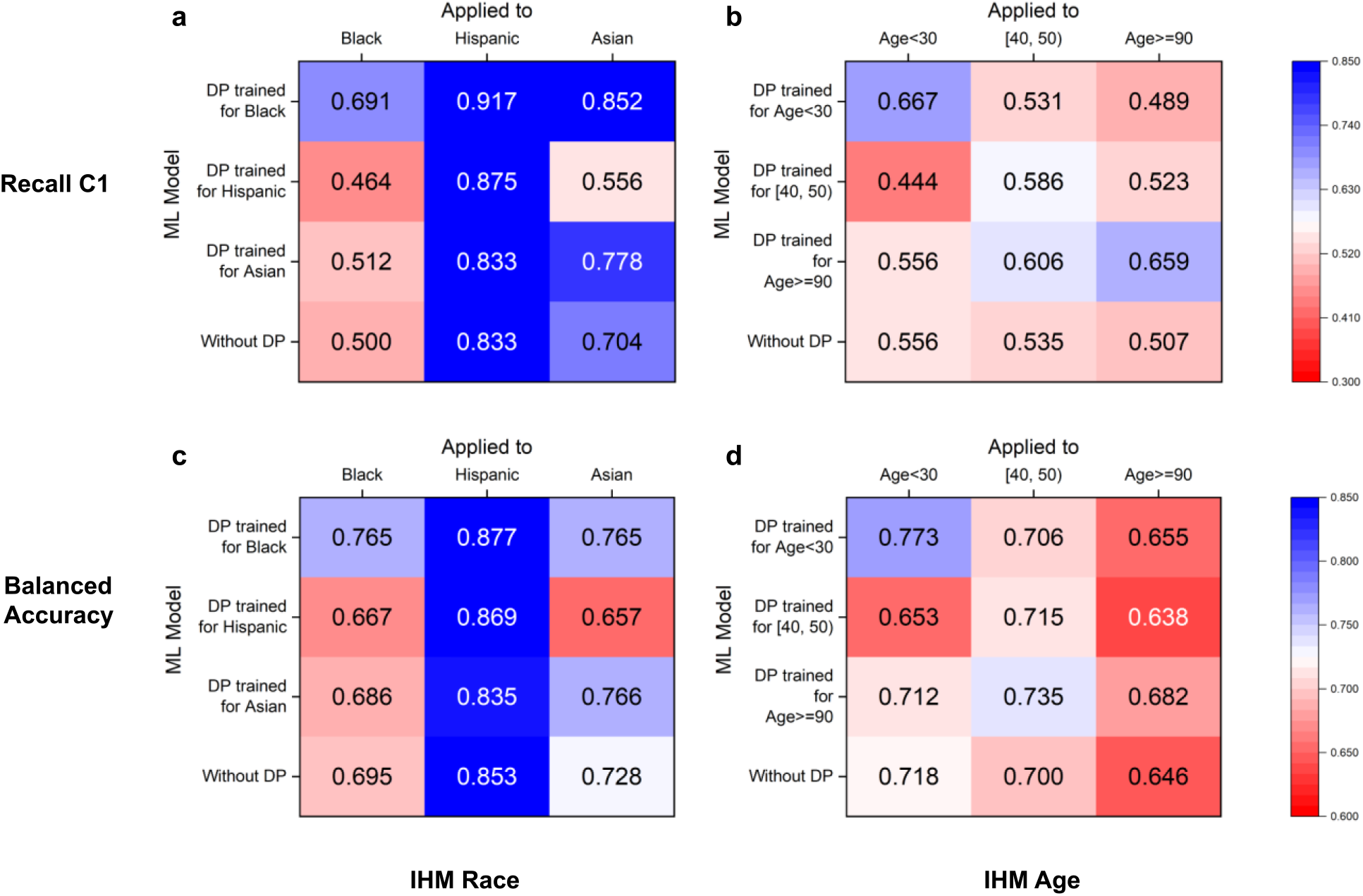
DP’s cross-group performance under various race and age settings for recall C1 and balanced accuracy for the IHM prediction. In subfigures, each row corresponds to a DP model trained for a specific subgroup. Each column represents a subgroup that a model is evaluated on. The values on the diagonal are the performance of a matching DP model, i.e., a DP model applied to the subgroup that it is designed for. The last rows show the group’s performance in the original model. To prevent overfitting, our method chooses optimal thresholds based on whole group performance. DP cross-group performance in terms of recall C1 for **(a)** race subgroups and **(b)** age subgroups for the IHM prediction. DP cross-group performance in terms of balanced accuracy for **(c)** race subgroups and **(d)** age subgroups for the IHM prediction.

**Supplementary Figure 5:**
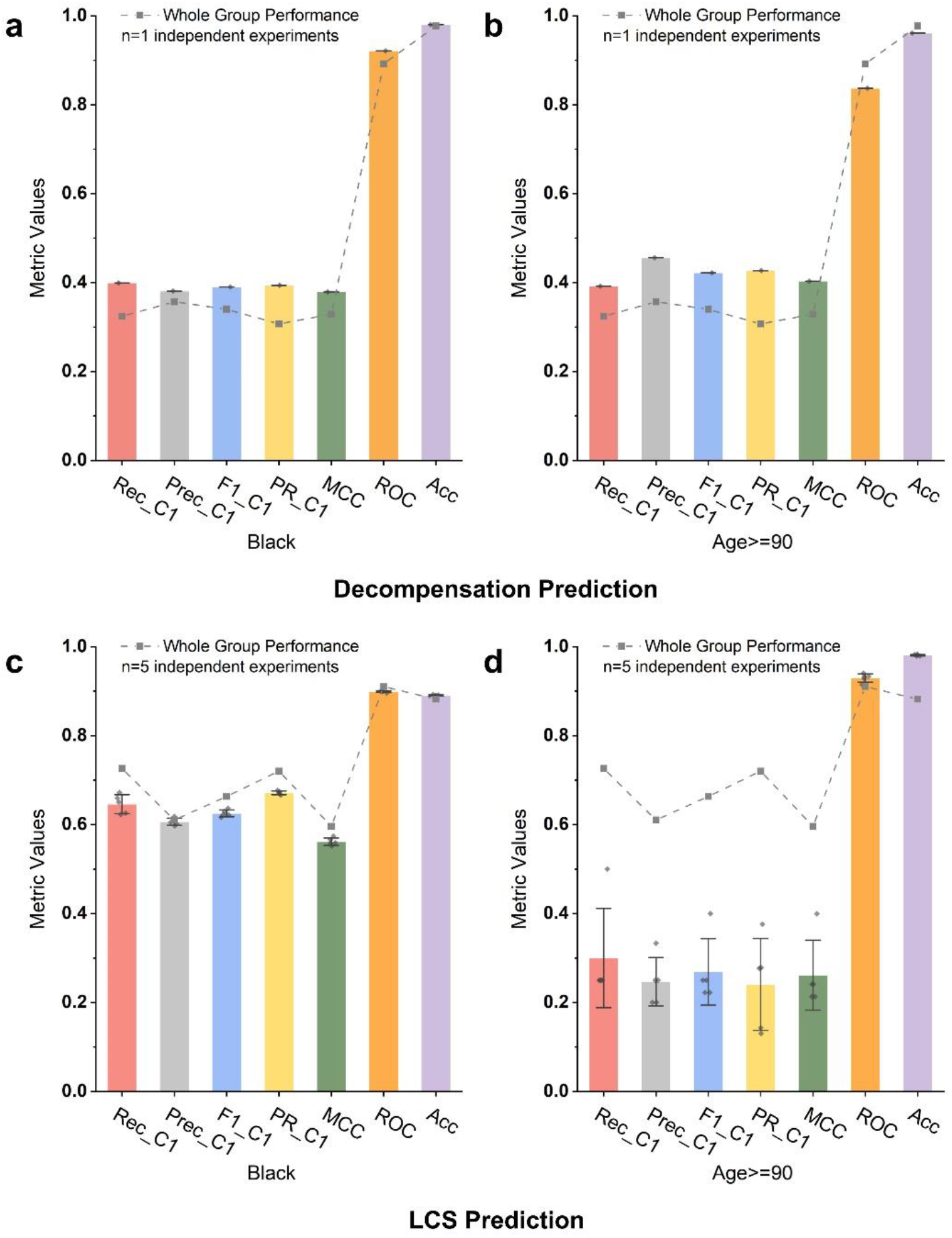
Comparison of whole-population metrics with minority-class-specific metrics. Some whole-population metrics (e.g., AUC ROC and accuracy) are misleading for the minority class. These deceptive metrics show high values, whereas the prediction is weak for the minority class. **(a)** Black subgroup performance for decompensation prediction. **(b)** Age 90+ subgroup performance for decompensation prediction. **(c)** Black subgroup performance for LCS prediction. **(d)** Age 90+ subgroup performance for LCS prediction. Due to the slow decompensation computation, each decompensation prediction is executed only once.

**Supplementary Figure 6:**
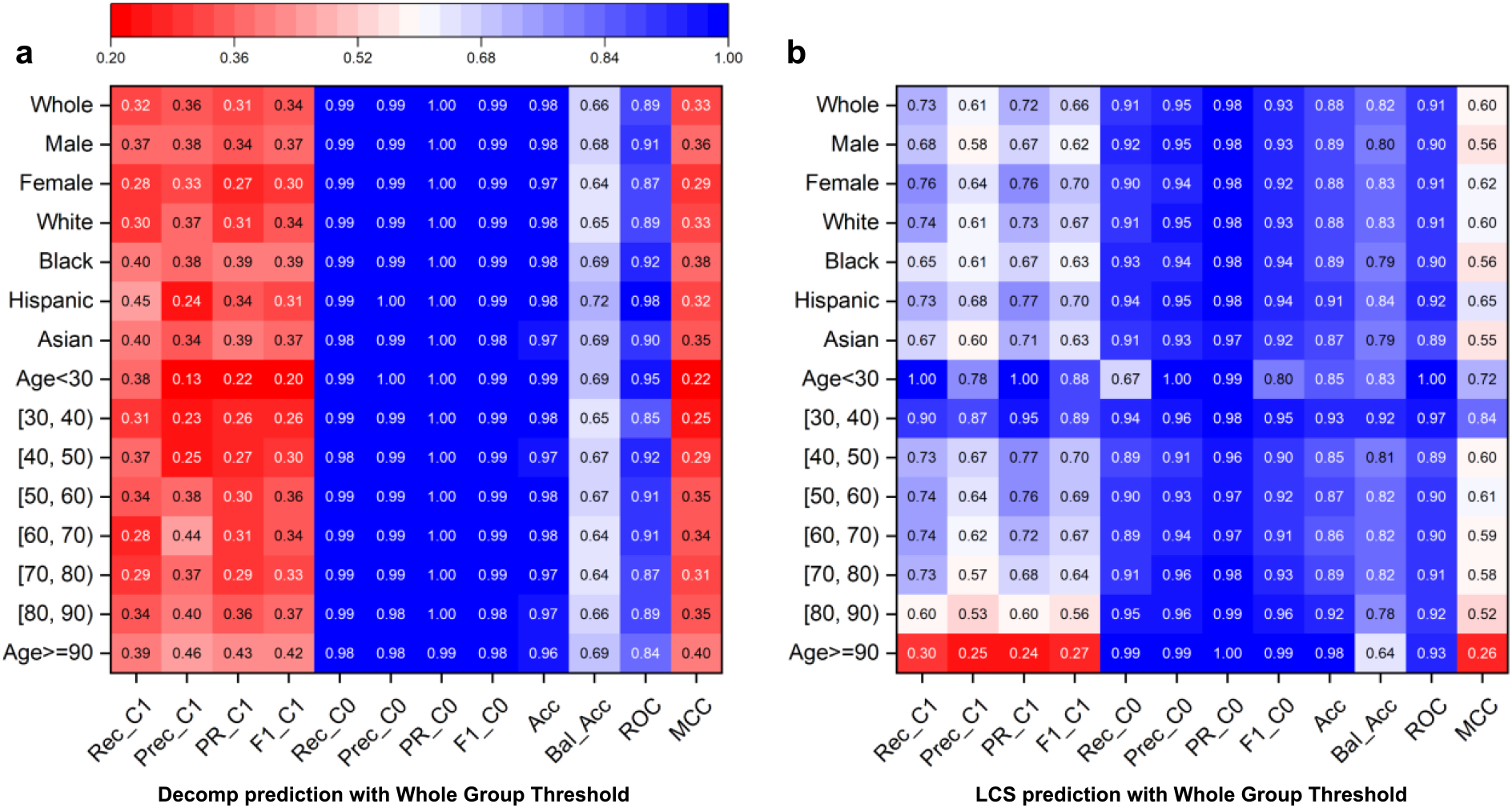
Prediction results under the original machine learning models (no bias correction) using one optimized threshold for all demographic groups. Rec_C1, Prec_C1, PR_C1, F1_C1, Rec_C0, Prec_C0, PR_C0, F1_C0, Acc, Bal_Acc, ROC, MCC stand for Recall Class 1, Precision Class 1, Area Under the Precision-Recall Curve Class 1, F1 score Class 1, Recall Class 0, Precision Class 0, Area Under the Precision-Recall Curve Class 0, F1 score Class 0, Accuracy, Balanced Accuracy, Area under the ROC Curve, Matthews Correlation Coefficient (MCC), respectively. **(a)** Prediction results for the decompensation prediction. The minority Class 1 represents patients whose health deteriorates after 24 hours. **(b)** Prediction results for the Lung cancer survivability (LCS) prediction. The minority Class 1 represents patients who survive lung cancer for at least 5 years after the diagnosis.

**Supplementary Figure 7:**
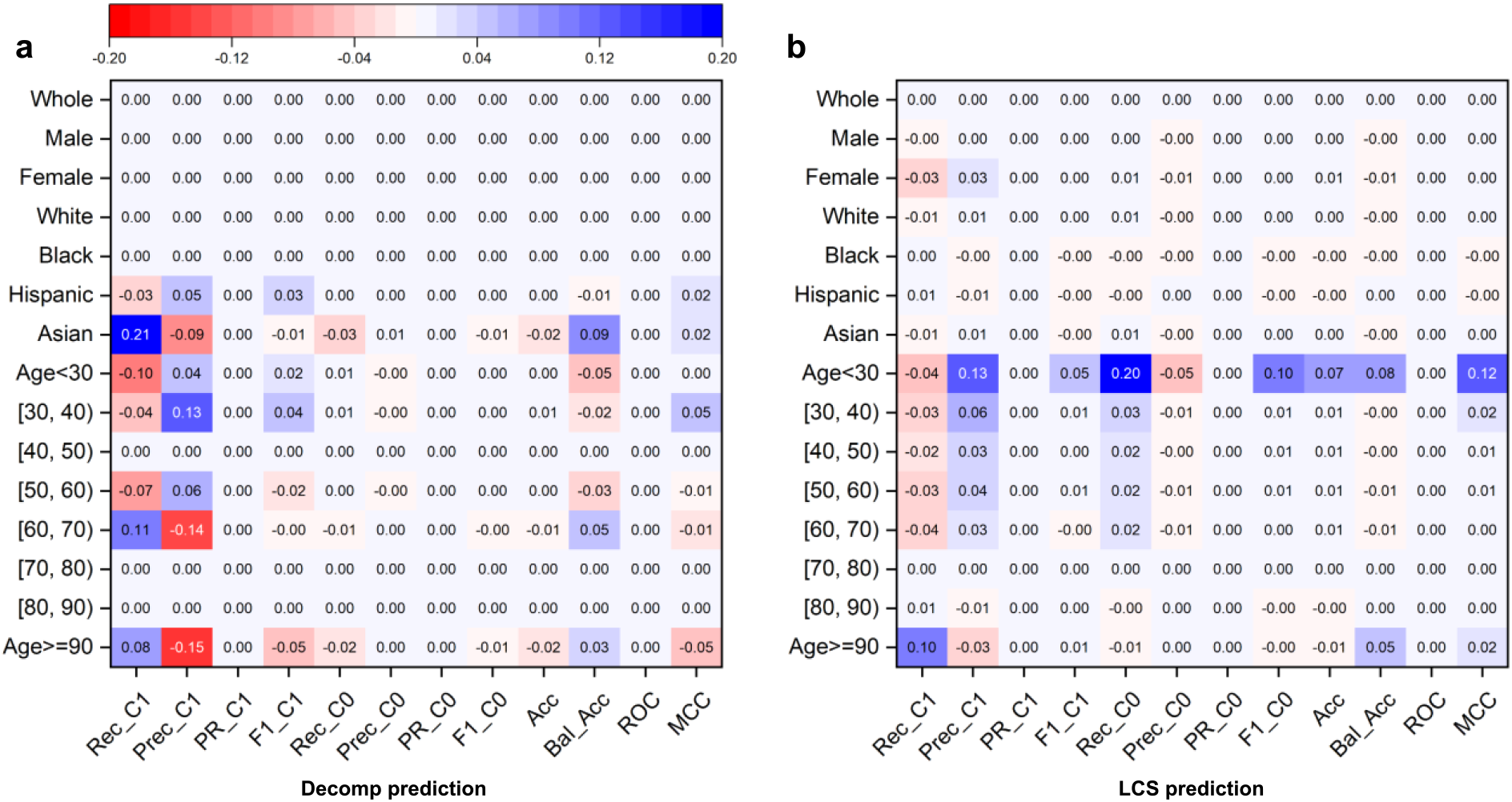
Differences in performance of the original machine learning models (no bias correction) using subgroup thresholds (i.e., different optimized thresholds for different demographic groups) and using the whole group threshold. Positive values mean that using a subgroup optimized threshold improves the performance. Rec_C1, Prec_C1, PR_C1, F1_C1, Rec_C0, Prec_C0, PR_C0, F1_C0, Acc, Bal_Acc, ROC, MCC stand for Recall Class 1, Precision Class 1, Area Under the Precision-Recall Curve Class 1, F1 score Class 1, Recall Class 0, Precision Class 0, Area Under the Precision-Recall Curve Class 0, F1 score Class 0, Accuracy, Balanced Accuracy, Area under the ROC Curve, and Matthews Correlation Coefficient (MCC), respectively. The performance differences between the two settings for **(a)** the decompensation prediction and **(b)** the LCS prediction.

**Supplementary Figure 8:**
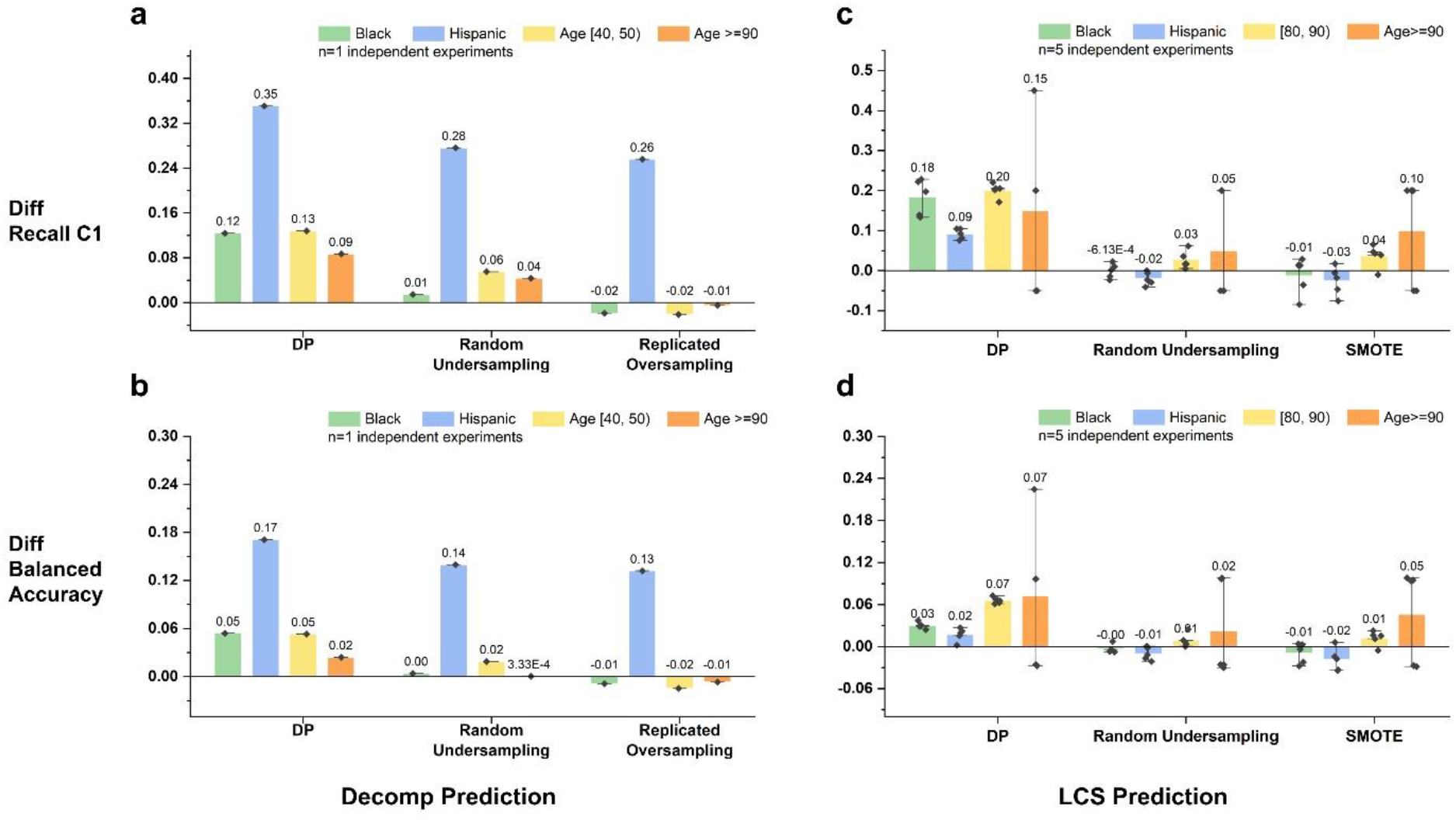
DP and two representative sampling techniques (random undersampling and replicated oversampling for Decomp and random undersampling and SMOTE for LCS) performance comparison over the original model for four demographic subgroups with poor original performance. Positive values indicate performance improvement, and negative values indicate performance degradation from the original model. The error bars represent the standard error of the experiment results. **(a)** In terms of recall C1 for Decomp prediction with the MIMIC III dataset. **(b)** In terms of balanced accuracy for Decomp prediction with the MIMIC III dataset. **(c)** In terms of recall C1 for the LCS prediction with the SEER dataset. **(d)** In terms of balanced accuracy for the LCS prediction with the SEER dataset. Due to the slow decompensation computation, each decompensation prediction is executed only once.

**Supplementary Figure 9:**
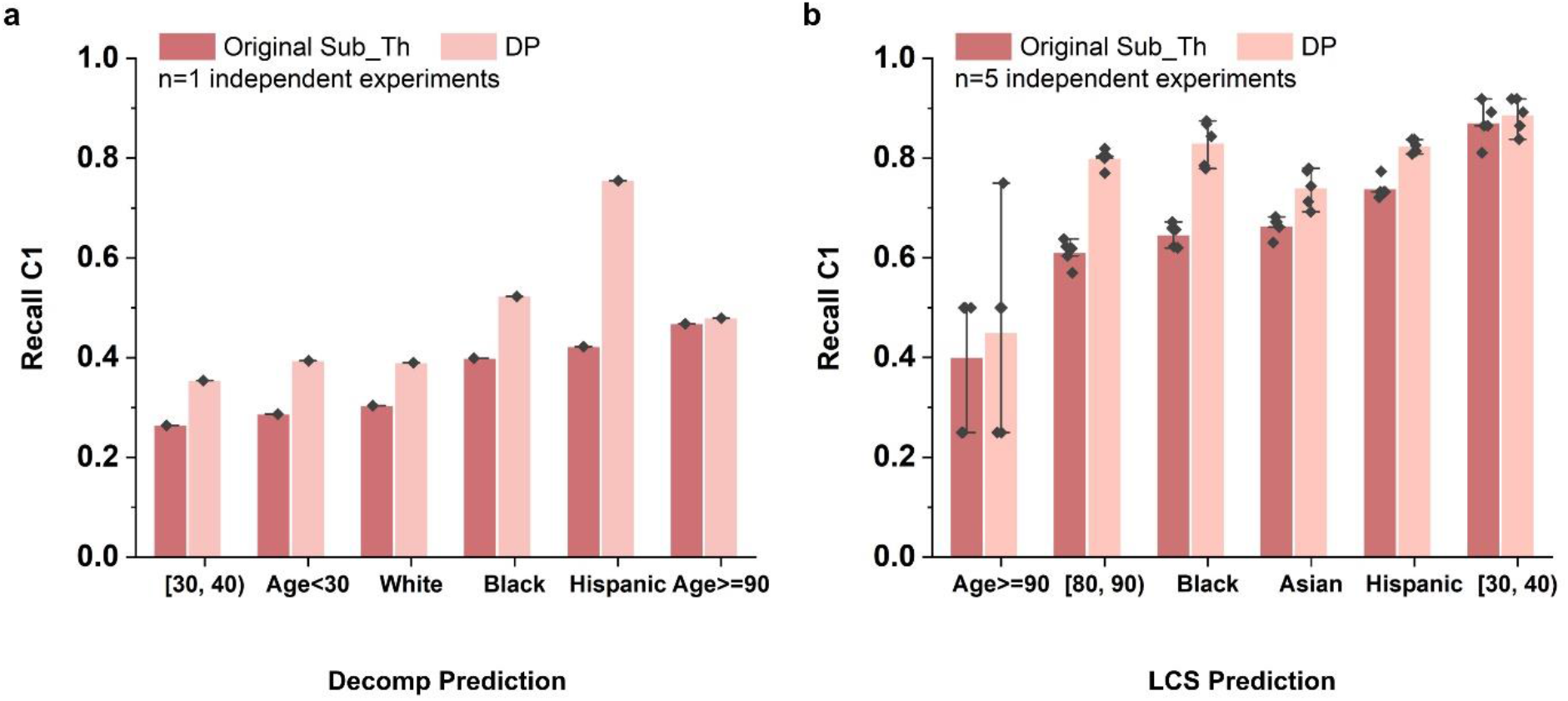
Performance of DP and subgroup-threshold-based original model in terms of minority class recall for decompensation prediction and 5-year lung cancer survivability (LCS) prediction. Darker red color represents the original model performance using subgroup optimized threshold and the lighter red color represents DP performance. The error bars represent the standard error of the experiment results. Model performance comparison for **(a)** Decomp prediction task and **(b)** LCS prediction task of 6 different racial or age subgroups. For the LCS task, the standard deviation values for DP are less than 0.04, with the exception of the age 90+ group (0.187). Due to the computation complexity, we only conducted the decompensation experiments once.

**Supplementary Figure 10:**
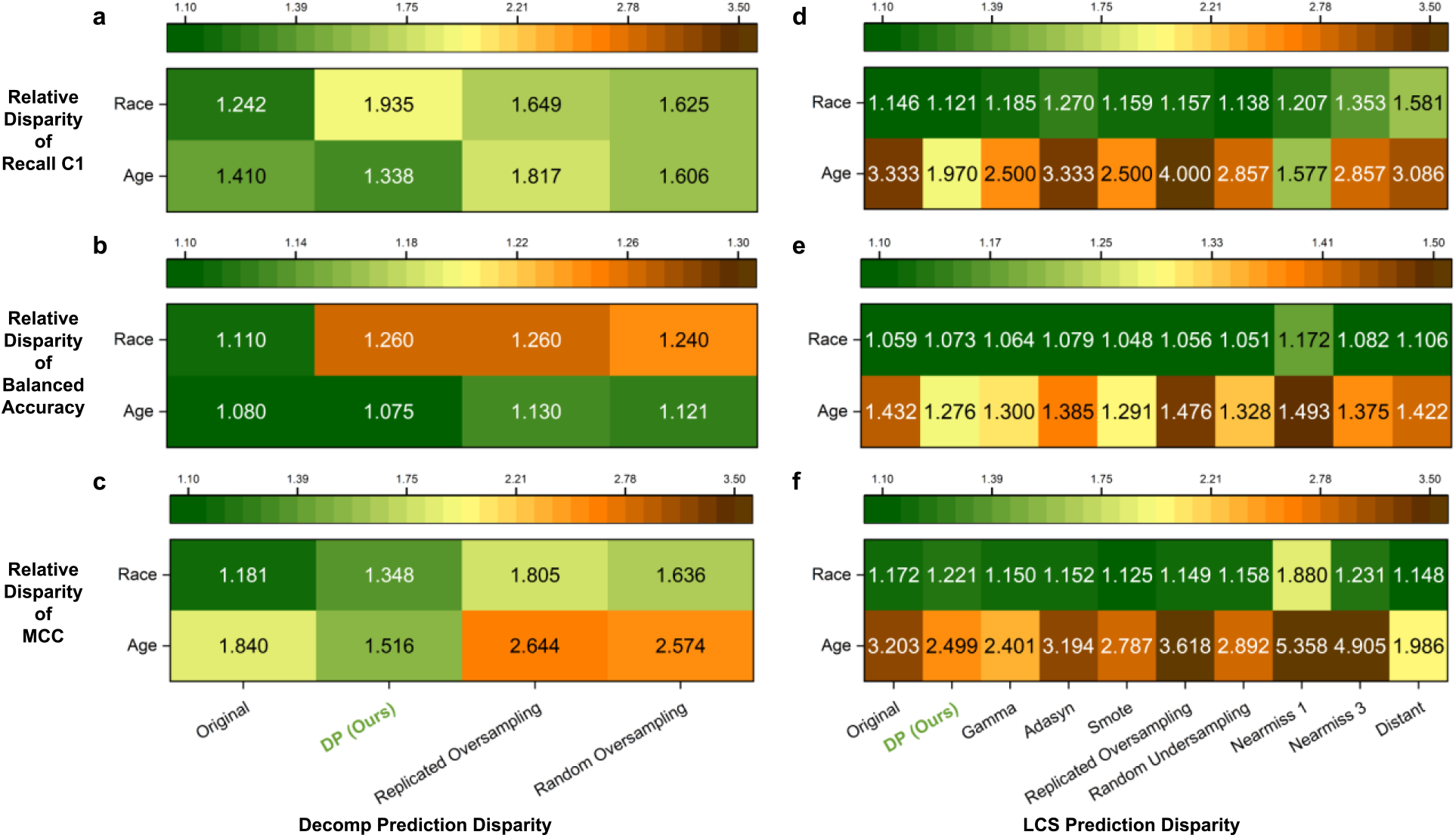
Relative disparity among racial and age groups under various sampling conditions, including DP and the original machine learning model without any bias correction. The relative disparity of MIMIC III Decomp prediction in terms of **(a)** minority class recall, **(b)** balanced accuracy, and **(c)** Matthews Correlation Coefficient (MCC). The relative disparity of SEER LCS prediction in terms of **(d)** minority class recall, **(e)** balanced accuracy, and **(f)** Matthews Correlation Coefficient (MCC).

**Supplementary Figure 11:**
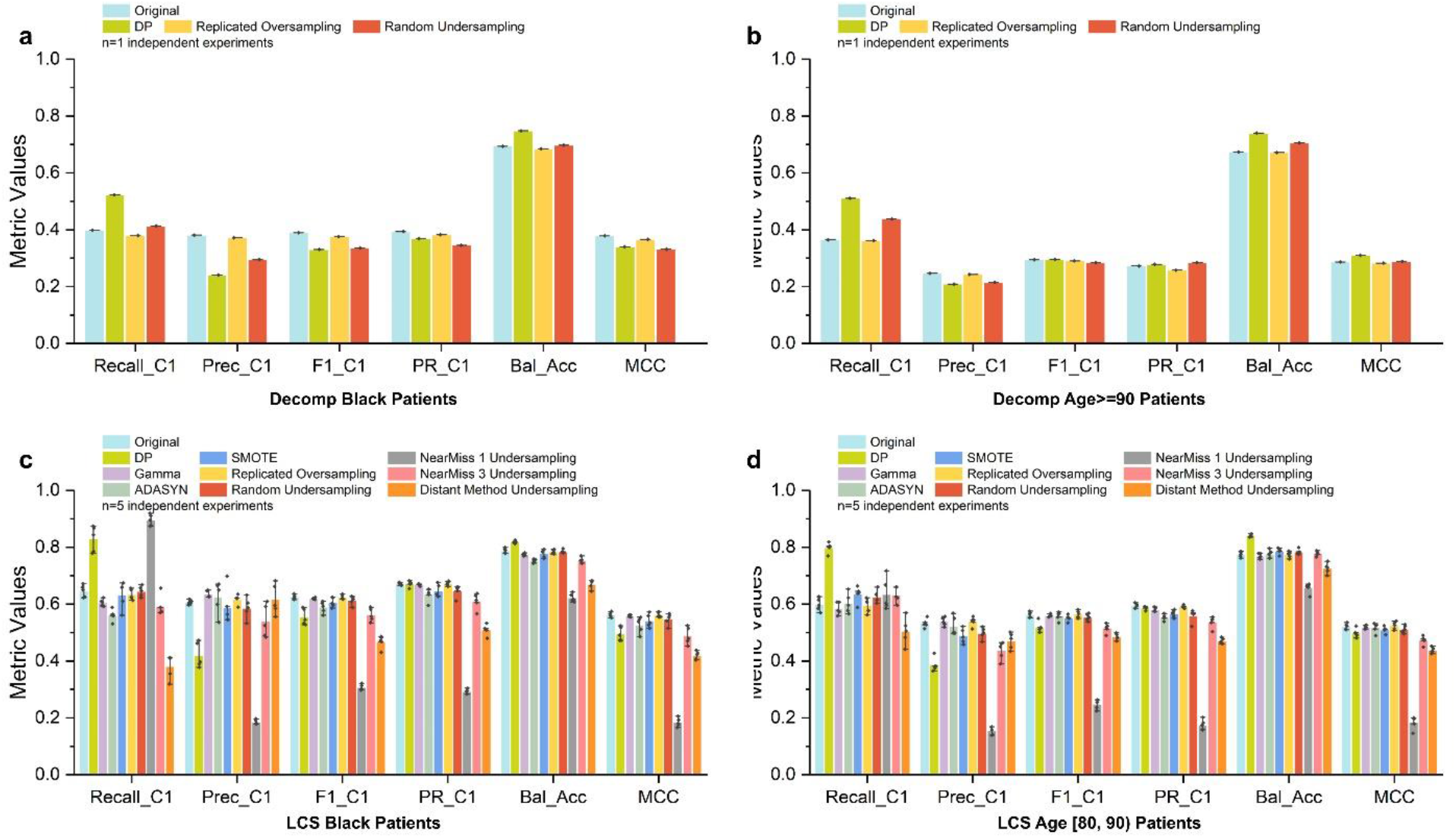
Decompensation prediction and 5-year lung cancer survivability (LCS) prediction under various sampling conditions, including DP and the original machine learning model without any sampling, in terms of minority class recall, precision, F1 score, AUC-PR, balanced accuracy, and Matthews Correlation Coefficient (MCC). The error bars represent the standard error of the experiment results. Prediction results from the original model and different sampling models for **(a)** Black patients and **(b)** age>=90 patients in the Decomp prediction with the MIMIC III dataset. Prediction results from the original model and different sampling models for **(c)** Black patients and **(d)** age [80, 90) patients in the LCS prediction with the SEER dataset. Due to the slow decompensation computation, each prediction is executed only once.

**Supplementary Figure 12:**
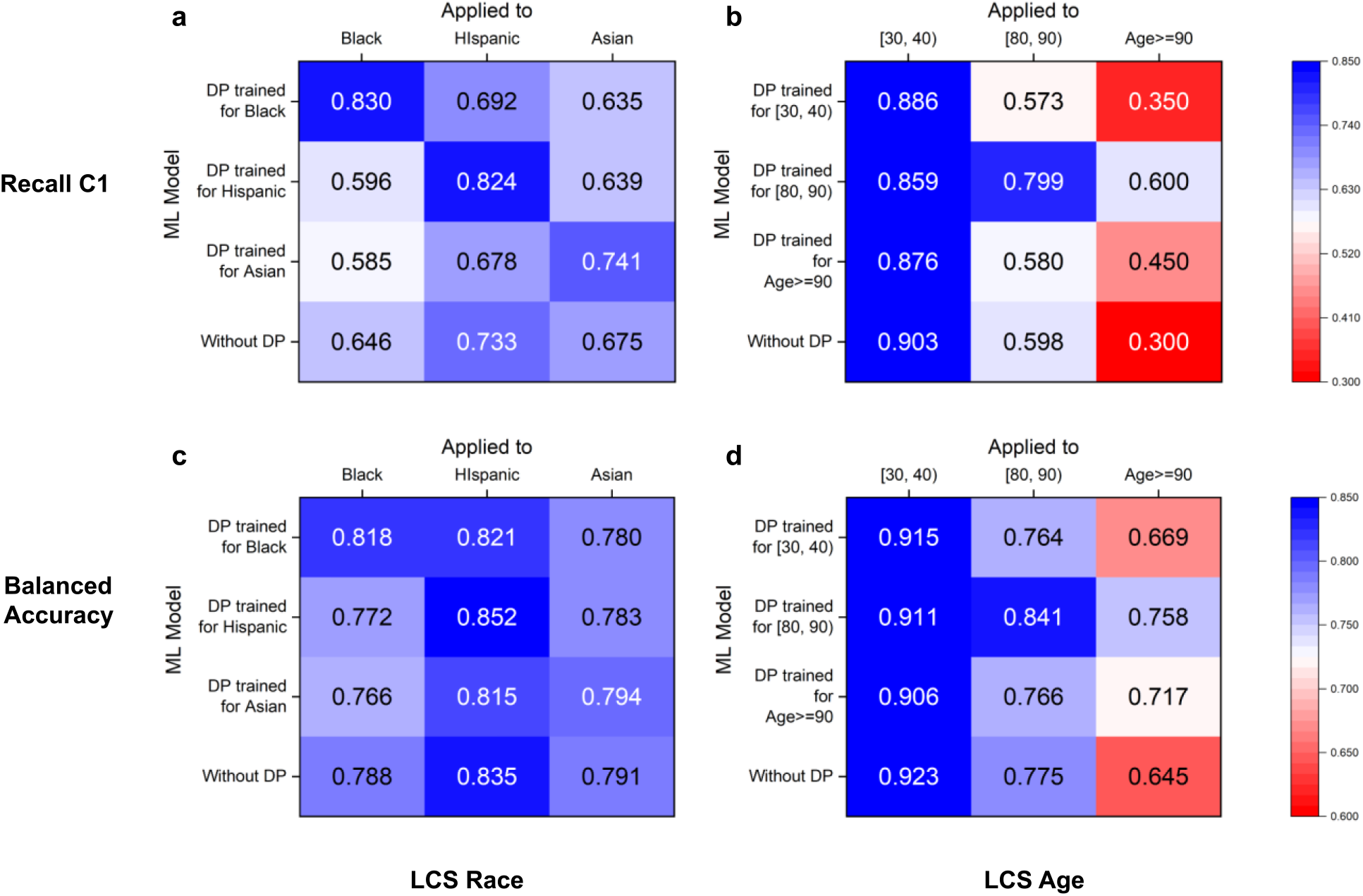
DP’s cross-group performance under various race and age settings for recall C1 and balanced accuracy for the LCS prediction. In subfigures, each row represents a model trained for a specific subgroup using DP. Each column represents a subgroup that a model is evaluated on. The values on the diagonal are the performance of a matching DP model, i.e., a DP model applied to the subgroup that it is designed for. The last rows show the group’s performance in the original model. To prevent overfitting, our method chooses optimal thresholds based on whole group performance. DP cross-group performance for **(a)** race subgroups and **(b)** age subgroups for the LCS prediction in terms of recall C1. DP cross-group performance for **(c)** race subgroups and **(d)** age subgroups for the LCS prediction in terms of balanced accuracy.

**Supplementary Figure 13:**
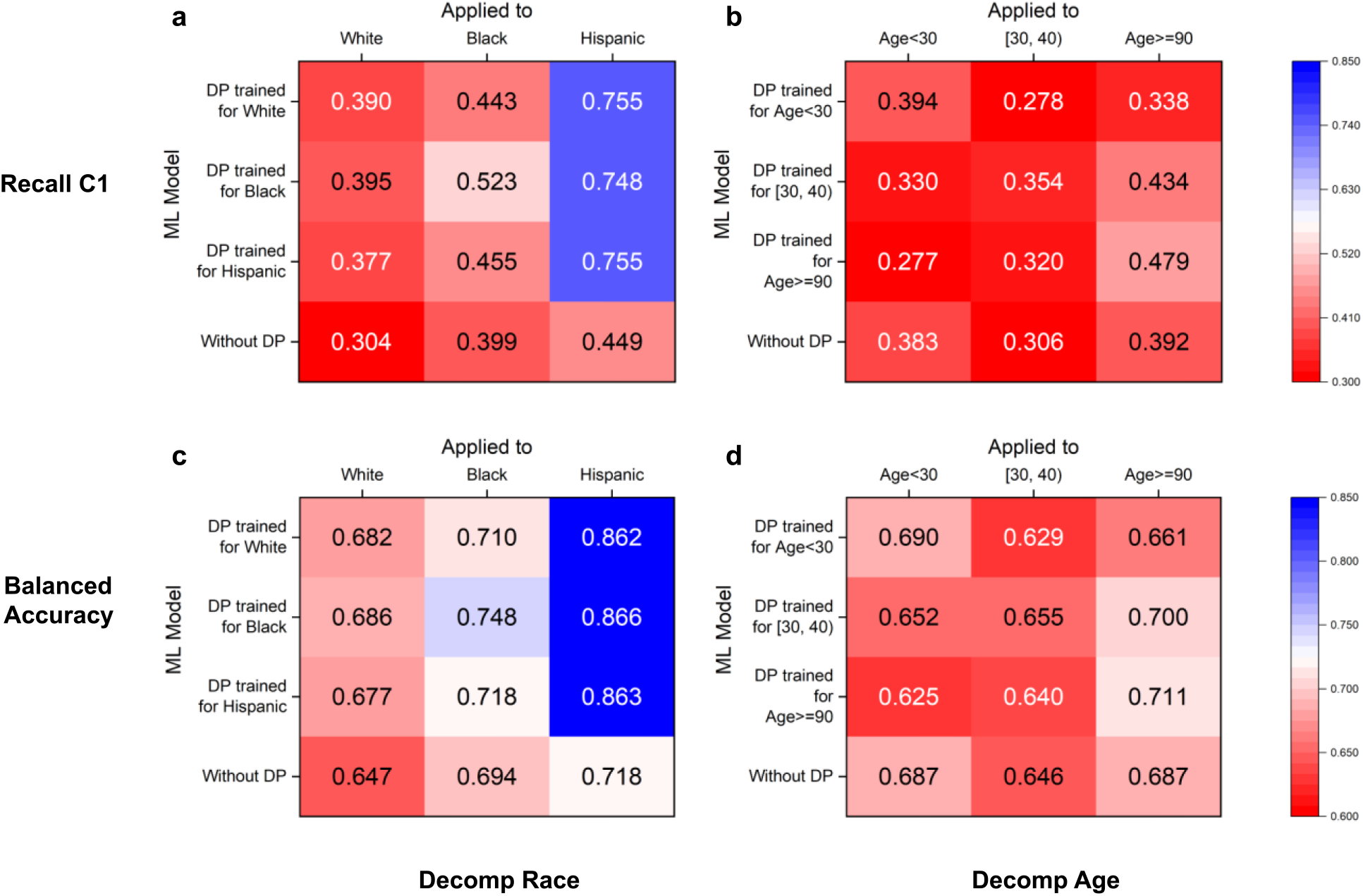
DP’s cross-group performance under various race and age settings for recall C1 and balanced accuracy for the decompensation prediction. In subfigures, each row corresponds to a DP model trained for a specific subgroup. Each column represents a subgroup that a model is evaluated on. The values on the diagonal are the performance of a matching DP model, i.e., a DP model applied to the subgroup that it is designed for. The last rows show the group’s performance in the original model. To prevent overfitting, our method chooses optimal thresholds based on whole group performance, as opposed to the (small) minority groups in the validation sets. DP cross-group performance for **(a)** race subgroups and **(b)** age subgroups for the decompensation prediction in terms of recall C1. DP cross-group performance for **(c)** race subgroups and **(d)** age subgroups for the decompensation prediction in terms of balanced accuracy.

**Supplementary Figure 14:**
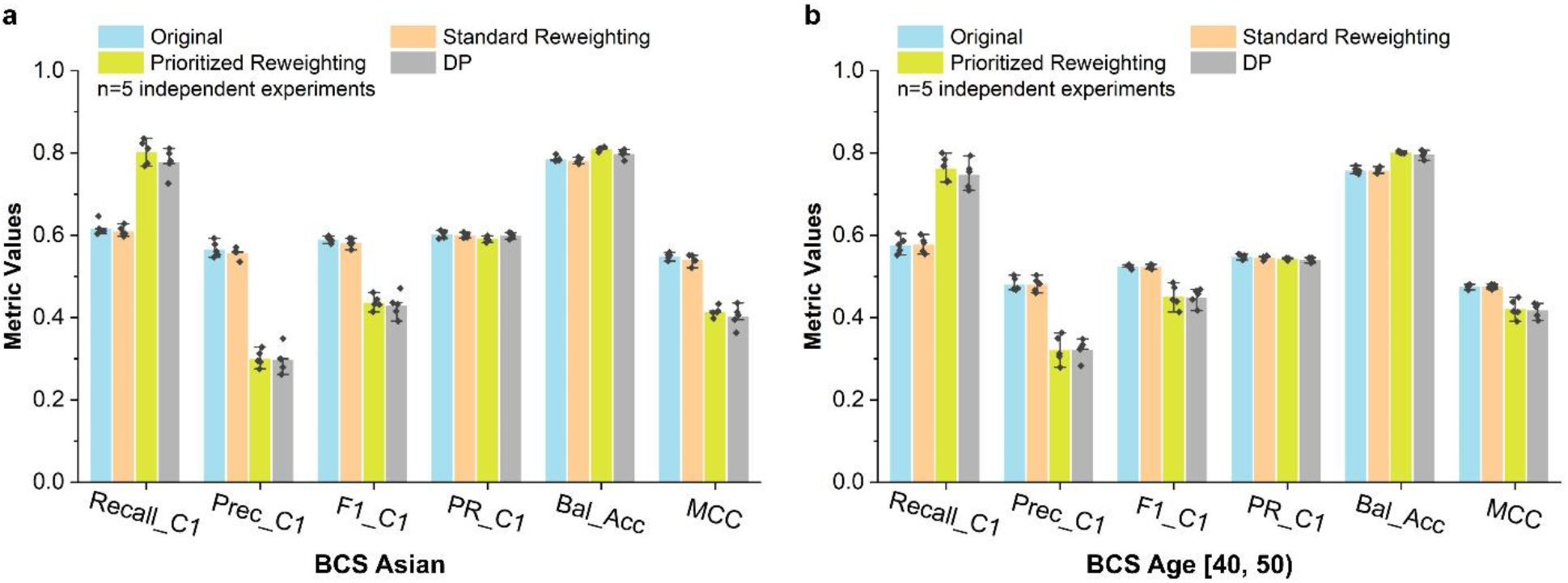
Performance comparison of the original model (without bias correction), standard reweighting, prioritized reweighting, and DP for (a) BCS Asian patients and (b) BCS [40, 50) patients. The error bars represent the standard error of the experiment results. In prioritized reweighting, we dynamically increase the weight of minority class (C1) samples of selected subgroups from 1 to 20 and select the best model using the same procedure as DP.

**Supplementary Figure 15:**
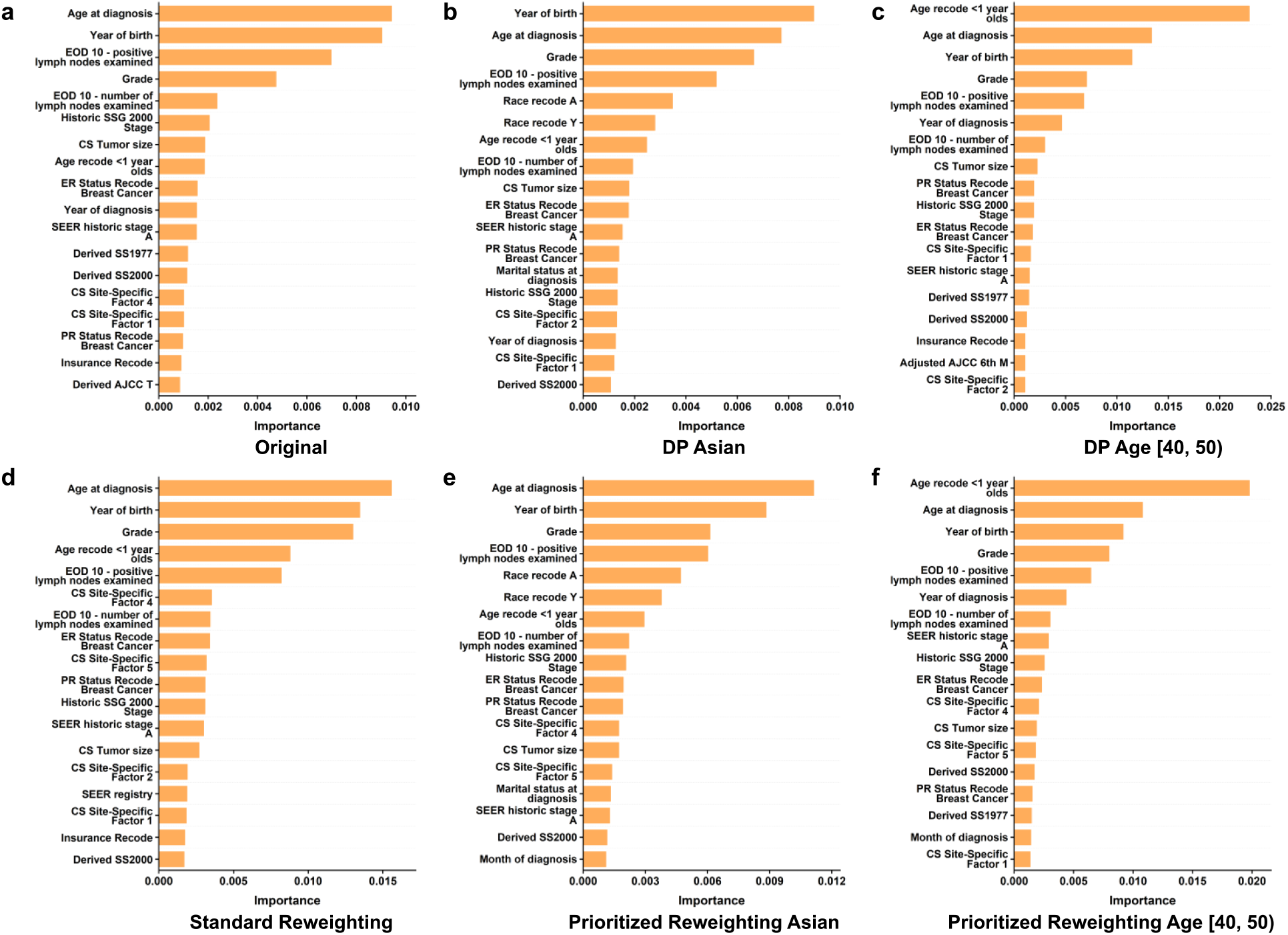
SHAP-avg feature importance of different BCS experiments. Original stands for the original machine learning model without any bias correction. DP stands for our Double Prioritized sampling method. Standard reweighting and prioritized reweighting are described in the Methods Section. In SHAP-avg, the SHAP importance of columns representing the same variable is averaged. The AJCC (American Joint Committee on Cancer) staging system is a system used to describe most types of cancer. SSG stands for the summary stage. ICD describes primary tumor site/type. PR and ER status represent a combination of a tumor marker and a site factor. Detailed variable and recode definitions can be found on the SEER website (https://seer.cancer.gov/data-software/documentation/seerstat/nov2016/). Feature importance for BCS prediction in **(a)** original model, **(b)** DP model for Asian patients, **(c)** DP model for age [40, 50) patients, **(d)** standard reweighting model, **(e)** prioritized reweighting model for Asian patients, and **(f)** prioritized reweighting model for age [40, 50) patients.

**Supplementary Figure 16:**
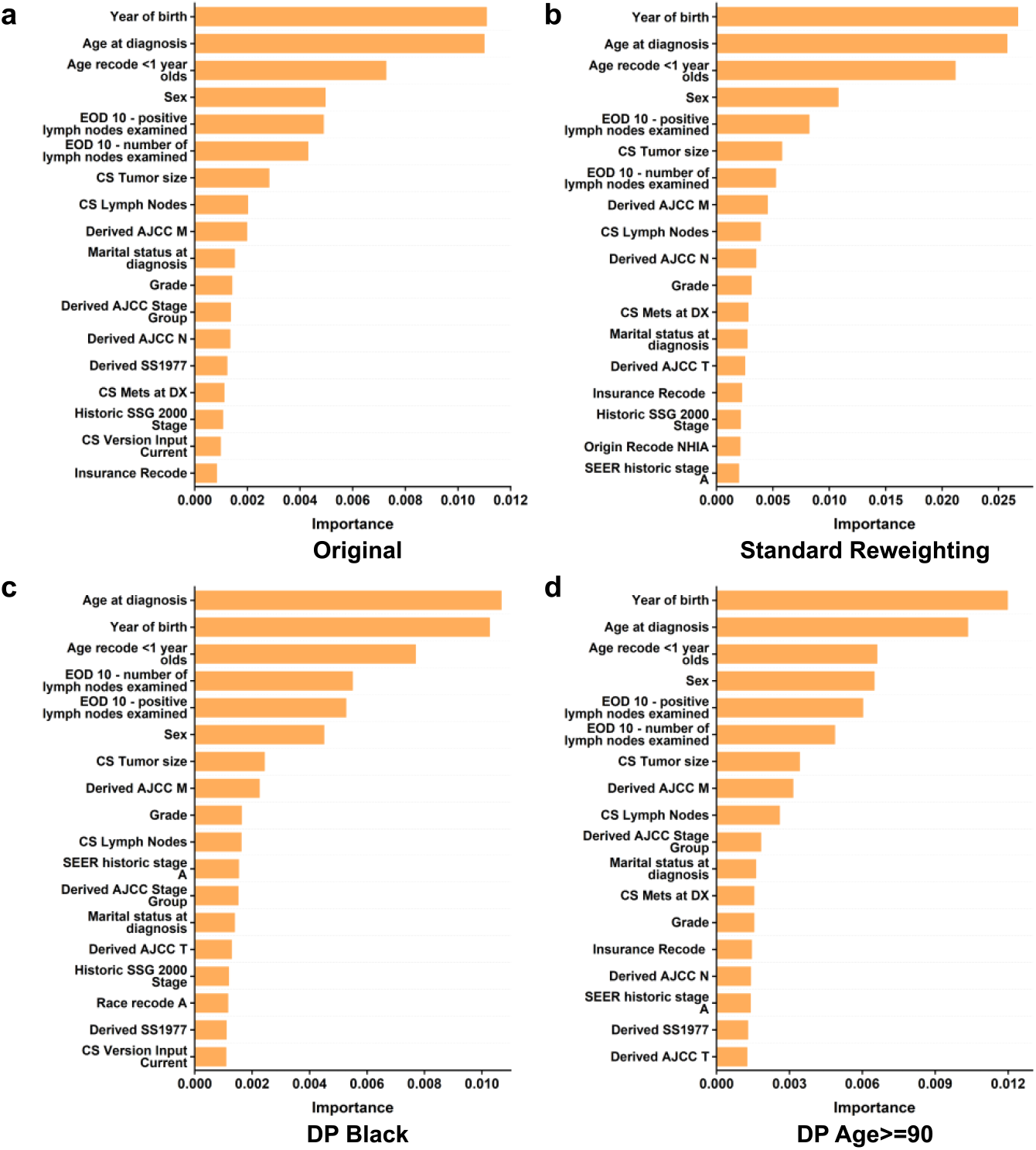
SHAP-avg feature importance of different LCS experiments. Original represents the original machine learning without any bias correction. DP stands for our Double Prioritized sampling method. Standard reweighting is described in the Methods section. In SHAP-avg, the importance of columns representing the same variable is averaged. The AJCC (American Joint Committee on Cancer) staging system is a system used to describe most types of cancer. SSG stands for the summary stage. ICD describes primary tumor site/type. CS Mets at DX provides information on distant metastasis, describing the extent of the disease. Detailed variable and recode definitions can be found on the SEER website (https://seer.cancer.gov/data-software/documentation/seerstat/nov2016/). Feature importance for LCS prediction in **(a)** original model, **(b)** standard reweighting model, **(c)** DP model for Black patients, and **(d)** DP model for age>=90 patients.

**Supplementary Figure 17:**
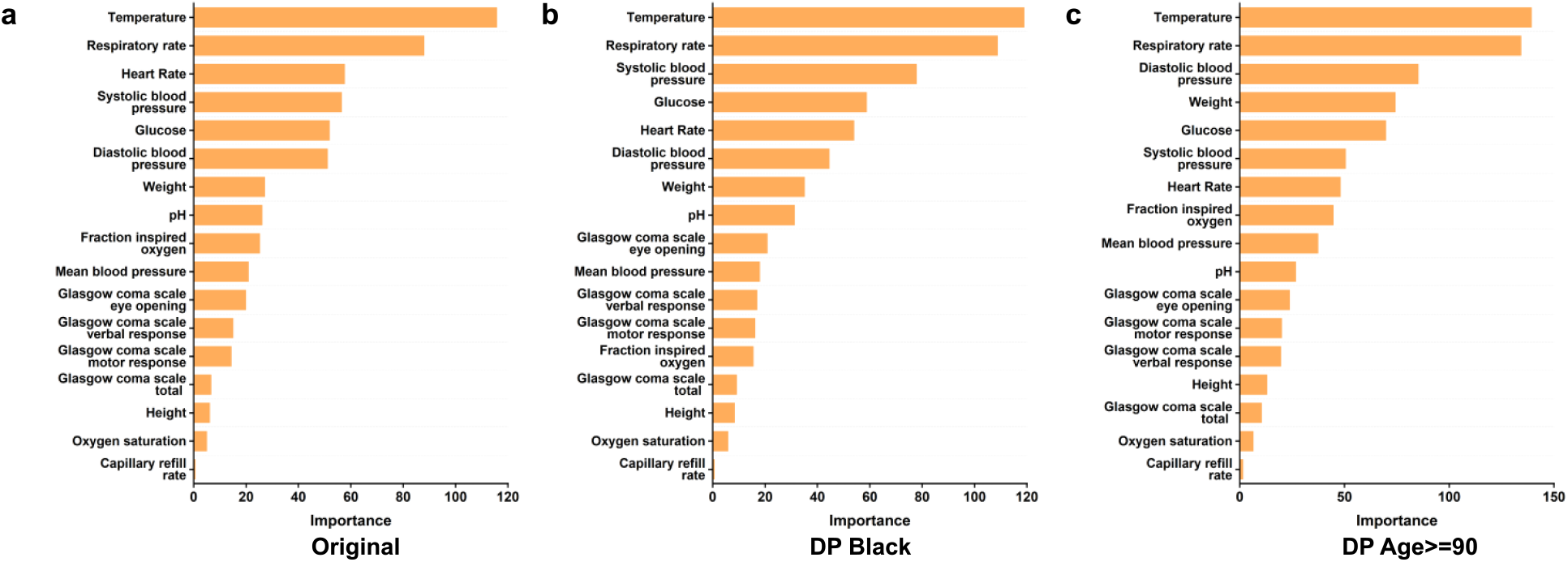
SHAP-avg feature importance of different IHM experiments. Original stands for the original machine learning model without any bias correction. DP stands for our Double Prioritized sampling method. In SHAP-avg, the importance of columns representing the same variable is averaged. Feature importance for IHM prediction in **(a)** original model, **(b)** DP model for Black patients, and **(c)** DP model for age>=90 patients.

**Supplementary Figure 18:**
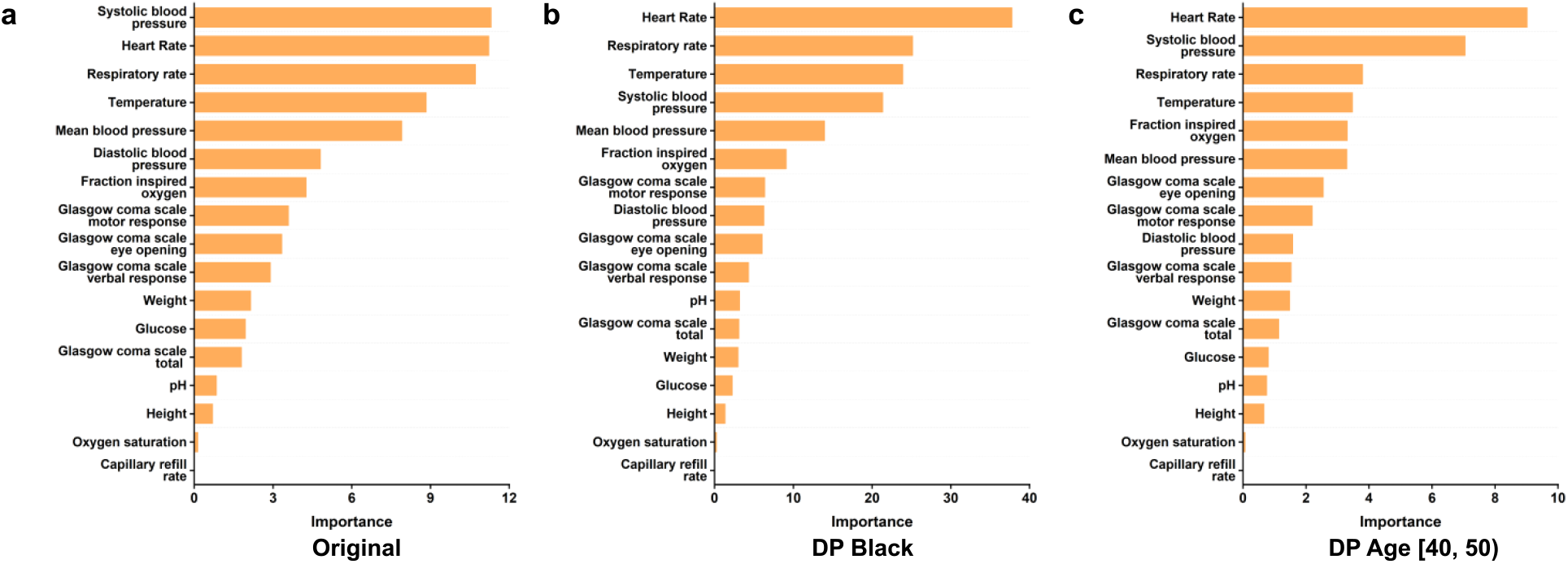
SHAP-avg feature importance of different decompensation experiments. Original stands for the original machine learning model without any bias correction. DP stands for our Double Prioritized sampling method. In SHAP-avg, the importance of columns representing the same variable is averaged. Feature importance for the decompensation prediction in **(a)** original model, **(b)** DP model for Black patients, and **(c)** DP model for age [40, 50) patients.

**Supplementary Figure 19:**
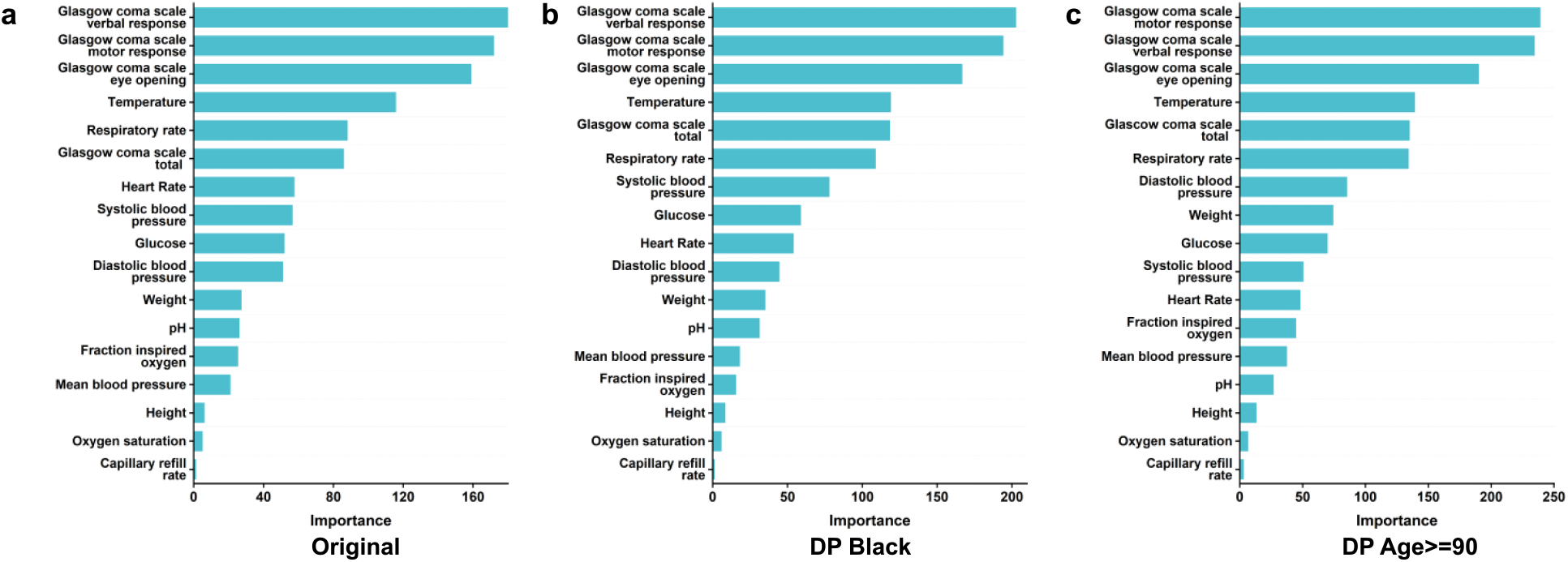
SHAP-sum feature importance of different IHM experiments. Original stands for the original machine learning model without any bias correction. DP stands for our Double Prioritized sampling method. In SHAP-sum, the importance of columns representing the same variable is summed up. Feature importance for the IHM prediction in **(a)** original model, **(b)** DP model for Black patients, and **(c)** DP model for age>=90 patients.

**Supplementary Figure 20:**
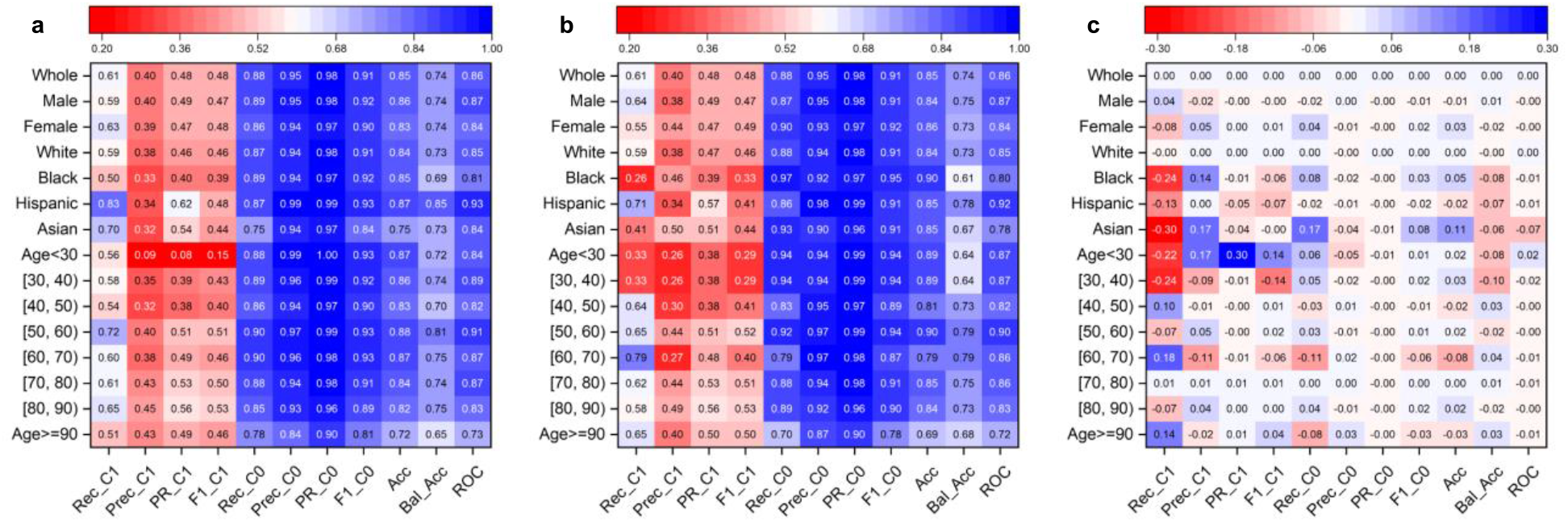
In-hospital mortality prediction performance of the original model with (a) whole group calibration, (b) subgroup calibration, and (c) difference in the performance between whole group and subgroup calibration. A positive value means subgroup calibration improves the performance. Rec_C1, Prec_C1, PR_C1, F1_C1, Rec_C0, Prec_C0, PR_C0, F1_C0, Acc, Bal_Acc, ROC stand for Recall Class 1, Precision Class 1, Area Under the Precision-Recall Curve Class 1, F1 score Class 1, Recall Class 0, Precision Class 0, Area Under the Precision-Recall Curve Class 0, F1 score Class 0, Accuracy, Balanced Accuracy, Area under the ROC Curve, respectively.

**Supplementary Table 1:** Learning parameters for four prediction models. BCS stands for breast cancer survivability. IHM stands for in-hospital mortality. LCS stands for lung cancer survivability. Decomp stands for decompensation. ANN stands for the artificial neural network.

| Learning Parameter | BCS Prediction | IHM Prediction | LCS Prediction | Decomp Prediction |
| --- | --- | --- | --- | --- |
| Hidden layers | (20, 20) | (16, 16) | (20, 20) | (128) |
| ANN | MLP | LSTM | MLP | LSTM |
| Learning Rate | 0.001 | 0.001 | 0.001 | 0.001 |
| Optimizer | adam | adam | adam | adam |
| Dropout | 0.1 | 0.3 | 0.1 | 0.0 |

For the IHM prediction task with MIMIC III datasets, training involves 100 epochs or stops early based on validation performance. For DP, we run for 50 epochs up to 20 additional units. For the Decomp prediction task with MIMIC III datasets, training involves 50 epochs or stops early based on validation performance. For DP experiments, we run for 10 epochs up to 20 additional units. The SEER cancer dataset is smaller, thus for the cancer prediction tasks, we run 25 epochs for all experiments. Each epoch produces a machine learning model; to choose the final model, we first identify the top three models based on balanced accuracy and then select the one with the highest precision-recall curve value of the minority class (denoted as PR\_C1). For the SEER dataset, 80% is used for training, 10% for validation, and 10% for testing. For MIMIC III, the percentages are 70% for training, 15% for validation, and 15% for testing.

**Supplementary Table 2:**
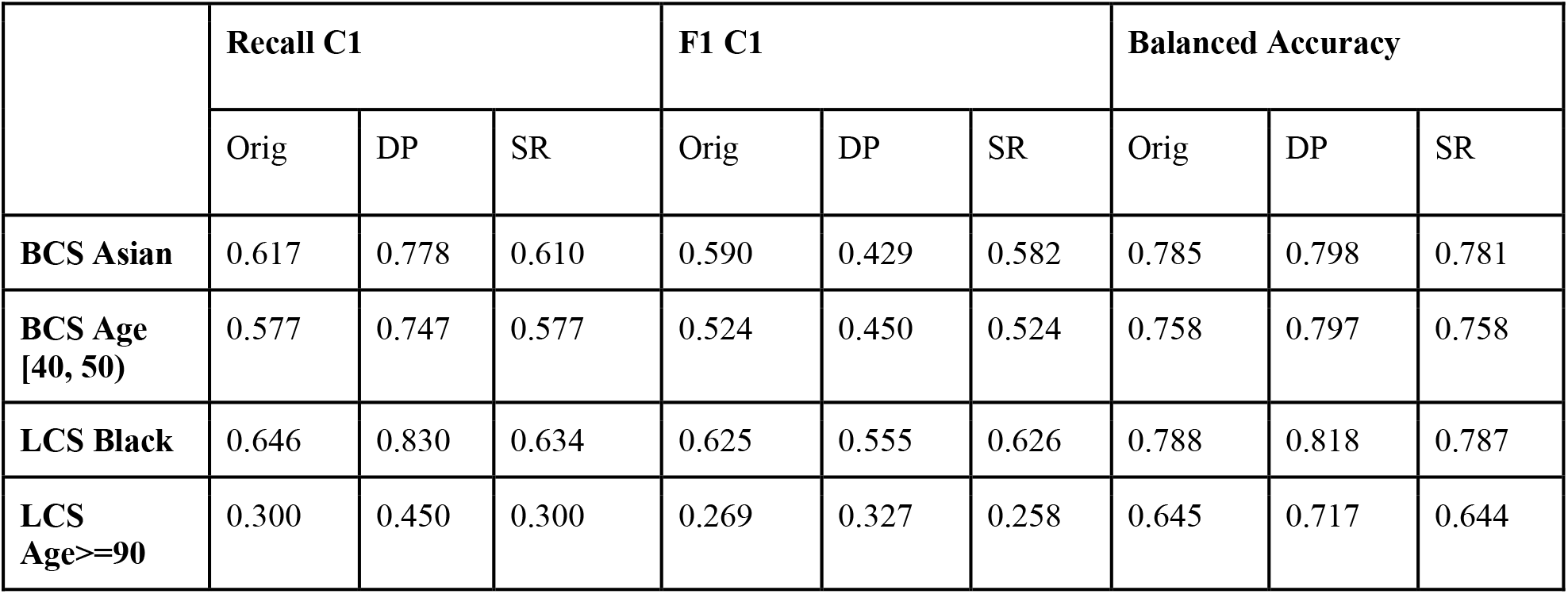
Performance comparison of standard reweighting with the original model and DP. Performance of the original model, applying DP, and applying standard reweighting for the BCS prediction and LCS prediction. For BCS, the minority class (C1) has a weight of 3.94 and the majority class (C0) has a weight of 0.57. For LCS, the minority class (C1) has a weight of 3.12 and the majority class (C0) has a weight of 0.60. Orig refers to the original model. SR stands for standard reweighting.

|  | Recall C1 |  |  | F1 C1 |  |  | Balanced Accuracy |  |  |
| --- | --- | --- | --- | --- | --- | --- | --- | --- | --- |
|  | Orig | DP | SR | Orig | DP | SR | Orig | DP | SR |
| **BCS Asian** | 0.617 | 0.778 | 0.610 | 0.590 | 0.429 | 0.582 | 0.785 | 0.798 | 0.781 |
| **BCS Age [40, 50)** | 0.577 | 0.747 | 0.577 | 0.524 | 0.450 | 0.524 | 0.758 | 0.797 | 0.758 |
| **LCS Black** | 0.646 | 0.830 | 0.634 | 0.625 | 0.555 | 0.626 | 0.788 | 0.818 | 0.787 |
| **LCS Age≥90** | 0.300 | 0.450 | 0.300 | 0.269 | 0.327 | 0.258 | 0.645 | 0.717 | 0.644 |

**Supplementary Table 3:** Summary of cross-race and cross-age-group results in the IHM, BCS, LCS, and Decomp tasks. A key case refers to that the matching DP models (i.e., sample enrichment matches the test group’s demographics) achieve the highest recall C1 performance.

| Task | No. of Key Cases | Race (No.) | Age Group (No.) | Figure Number |
| --- | --- | --- | --- | --- |
| **IHM** | 3 (out of 6) | Black (1) | <30, 90+ (2) | Supp. Fig. 4 |
| **BCS** | 5 (out of 6) | Black, Hispanic, Asian (3) | <30, [30, 40) (2) | Fig. 8 |
| **LCS** | 4 (out of 6) | Black, Hispanic, Asian (3) | [80, 90) (1) | Supp. Fig. 12 |
| **Decomp** | 4 (out of 6) | Black (1) | <30, [30, 40), 90+ (3) | Supp. Fig. 13 |
| **Total** | **16 (Out of 24)** | **8** | **8** | -- |

**Supplementary Table 4:** Performance of MLP models using different structures. The performance of MLP models on the BCS and LCS tasks. We evaluate 3 different numbers of layers, 3 different numbers of neurons per layer, and 3 different dropout rates, generating 27 models in total for each task. The results are comparable among the models. The table shows the subgroup performance of the default model (2 layers with 20 neurons, 0.1 dropout rate) compared with two other models (5 layers with 30 neurons, 0.2 dropout rate and 10 layers with 50 neurons, 0.3 dropout rate).

|  | Recall C1 |  |  | F1 C1 |  |  | Balanced Accuracy |  |  |
| --- | --- | --- | --- | --- | --- | --- | --- | --- | --- |
|  | 2-20-0.1<br>(default) | 5-30-0.2 | 10-50-0.3 | 2-20-0.1<br>(default) | 5-30-0.2 | 10-50-0.3 | 2-20-0.1<br>(default) | 5-30-0.2 | 10-50-0.3 |
| **BCS Asian** | 0.617 | 0.627 | 0.643 | 0.590 | 0.584 | 0.591 | 0.785 | 0.788 | 0.795 |
| **BCS Age [40, 50)** | 0.577 | 0.571 | 0.607 | 0.524 | 0.518 | 0.514 | 0.758 | 0.755 | 0.767 |
| **LCS Black** | 0.646 | 0.644 | 0.653 | 0.625 | 0.622 | 0.631 | 0.788 | 0.787 | 0.792 |
| **LCS Age≥90** | 0.300 | 0.250 | 0.300 | 0.269 | 0.242 | 0.310 | 0.645 | 0.620 | 0.646 |

**Supplementary Table 5:**
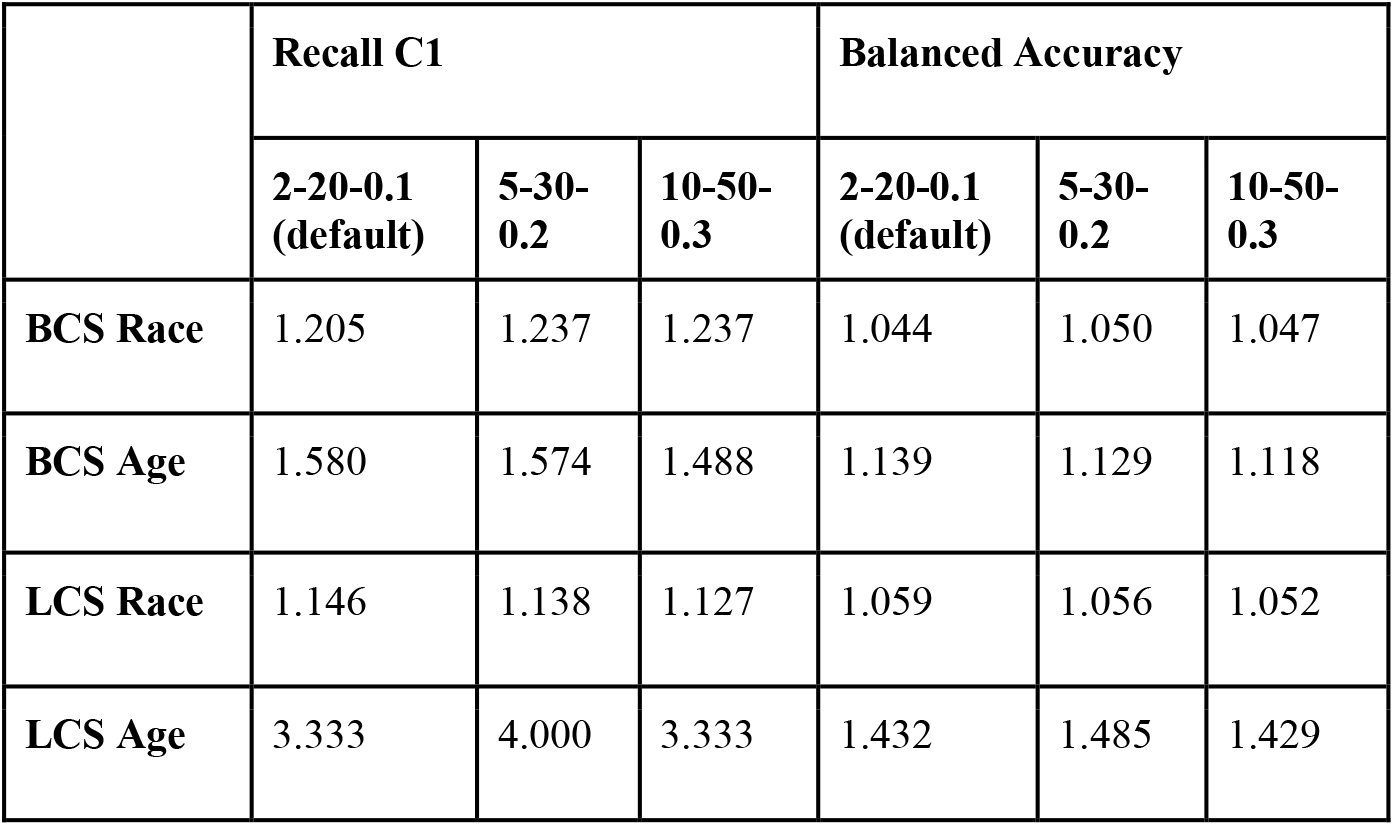
Relative disparity of MLP models using different structures. The relative disparity among subgroups for the BCS and LCS tasks are shown, including the disparity of the default model (2 layers with 20 neurons, 0.1 dropout rate) compared with two other models (5 layers with 30 neurons, 0.2 dropout rate and 10 layers with 50 neurons, 0.3 dropout rate).

